# Lymphatic Dysplasia Shapes Liver Epithelial Outcomes in Biliary Atresia

**DOI:** 10.64898/2026.10.12.26365253

**Authors:** Leonie Schumm, Robert L. Chua, Matylda Z. Kuzinska, Mijuna Meindl, Susanna Quach, Julian Weihs, Dominic Gogol, Marius Zschiesche, Reinhild Duennebacke, Marie Uecker, Rose Behncke, Alexander Sudy, Johannes Liebig, Jan C. Kamp, Martin Kucklick, Brigitta Globke, Robert Öllinger, Joachim Kübler, Stephan Henning, Christoph Slavetinsky, Susanne Engelmann, René Hägerling, Omid Madadi-Sanjani, Roland Eils, Adrien Guillot, Philip Bufler, Christian Conrad, Milad Rezvani

**Affiliations:** Charité – Universitätsmedizin Berlin, Department of Pediatric Gastroenterology, Nephrology and Metabolic Medicine, Berlin, Germany; Berlin Institute of Health (BIH) at Charité-Universitätsmedizin Berlin, Clinician Scientist Program, Berlin, Germany; Center for Digital Health, Berlin Institute of Health (BIH) at Charite - Universitätsmedizin Berlin, Berlin, Germany; Berlin School for Regenerative Therapies (BSRT) at the Berlin Center for Regenerative Therapies (BCRT), Berlin, Germany; Hannover Medical School, Department of Pediatric Surgery, Hannover, Germany; Berlin Institute of Health (BIH) at Charité-Universitätsmedizin Berlin, BIH Center for Regenerative Therapies (BCRT), Berlin, Germany; Hannover Medical School, Department of Respiratory Medicine and Infectious Diseases, Hannover, Germany; Member of the German Center for Lung Research (DZL), Hannover, Germany, Biomedical Research in Endstage and Obstructive Lung Disease Hannover (BREATH); Institute for Microbiology, Technische Universität Braunschweig, Braunschweig 38106, Germany; Microbial Proteomics, Helmholtz Centre for Infection Research (HZI), Braunschweig 38124, Germany; Charité – Universitätsmedizin Berlin, Department of Surgery, Berlin, Germany; Department of General Surgery, Division of Transplantation, General Hospital of Vienna, Medical University of Vienna; Department of Clinic for Pediatric Surgery and Pediatric Urology, Klinikum Bremen-Mitte, Bremen, Germany; Department of Pediatric Surgery and Pediatric Urology, University Children’s Hospital Tübingen, Tübingen, Germany; Cluster of Excellence EXC 2124 Controlling Microbes to Fight Infections, Tübingen, Germany; Lymphovascular Medicine Laboratory, Berlin Institute of Health (BIH) at Charité - Universitätsmedizin Berlin, Berlin, Germany; Spatial Diagnostics Platform, Berlin Institute of Health (BIH) at Charité -Universitätsmedizin Berlin, Berlin, Germany; Department of Pediatric Surgery, University Medical Center Hamburg-Eppendorf, Hamburg, Germany; Department of Transplant Surgery, University Medical Center Hamburg-Eppendorf, Hamburg, Germany; Department of Mathematics and Computer Science, Freie Universität Berlin, Berlin, Germany; Health Data Science Unit, Heidelberg University Hospital and BioQuant, University of Heidelberg, Heidelberg, Germany; Intelligent Medicine Institute, Fudan University, Shanghai, China, Shanghai, China; Charité – Universitätsmedizin Berlin, Department of Hepatology & Gastroenterology, Berlin, Germany; German Center for Child and Adolescent Health (DZKJ), partner site Berlin, Berlin, Germany; Division of Pediatric Gastroenterology, Hepatology and Nutrition, Cincinnati Children’s Hospital Medical Center, OH, US; Der Simulierte Mensch (Si-M), Joint Research Center of Technische Universität Berlin and Charité - Universitätsmedizin Berlin, Berlin, Germany

**Author notes:** Contributed equally.

## Abstract

Biliary Atresia (BA) is the leading cause of pediatric liver transplantation. Early mechanisms governing outcomes remain unknown. A multiscale atlas of outcome-stratified BA, resolving single cell, spatial and 3D-tissue states of livers sampled at Kasai portoenterostomy (KPE), established lymphatic dysplasia in infants on a trajectory toward liver failure and transplantation (‘severe BA’). An architecturally fragmented, immature and CCL21^high^ lymphatic vasculature marked severe BA early on. In the epithelium, hepatocyte-to-biliary plasticity emerged across clinical outcomes, while hepatocyte dedifferentiation, acute-phase-reactions and *MMP7*/*SOX9*^high^ cholangiocyte expansion delineated severe disease. *Ex vivo*, lymphatic endothelial cells, isolated from severe BA livers, induced *SOX9*^high^ epithelial progenitors when cultured as assembloids with cholangiocytes. MMP7, the known BA mediator, was governed by ectopic CCL21-signaling on epithelium after inflammatory priming, as evidenced in CCL21 neutralization experiments. In sum, lymphatic endothelial dysplasia is an early phenomenon in BA that perturbs intrahepatic epithelial states and offers CCL21 as a target for epithelial stability.

## INTRODUCTION

Biliary atresia (BA) is a progressive neonatal cholestatic disease and the most common indication for pediatric liver transplantation.(*1–3*) After diagnosis, BA requires prompt surgical intervention. Yet outcomes following palliative Kasai portoenterostomy (KPE) remain variable.(*1, 2*) Despite progress in screening and diagnosis,(*4, 5*) the early pathophysiological determinants of BA native liver survival or failure remain elusive. This has deep implications for anticipatory care and target identification. Understanding the drivers of intrahepatic outcome early on is vital because KPE removes the obliterated extrahepatic bile duct remnant but leaves the intrahepatic disease process largely untouched. Despite important efforts in single-cell resolution studies of BA,(*6, 7*) there has been no comprehensive spatiotemporal BA atlas that links early molecular phenomena to concrete clinical outcome stratifications.

In BA, periportal hepatocytes and cholangiocytes undergo a continuum of reactive epithelial states.(*8*) As part of the so-called ‘ductular reaction’, CK19+ ductules expand into the parenchyma, largely without obvious bile-draining capacity. Matrix metalloproteinase (MMP)-7, induced by Wnt/β-catenin signaling, drives cholangiocyte injury and fibrosis in BA and also serves as a validated diagnostic biomarker.(*4, 9, 10*)

While liver regeneration in many injury settings relies primarily on hepatocyte self-duplication without a classical stem cell,(*11*) Wnt signaling-linked LGR5+ progenitors, as found in the intestine,(*12*) can – in principle – also emerge in the injured liver.(*13*) Transdifferentiation between hepatocyte and cholangiocyte without stem or progenitor cell intermediates is well-established in the liver and has been demonstrated in different systems, including a NOTCH-deficient mouse model of Alagille syndrome(*14*) and even in human Metabolic Dysfunction-Associated Steatotic Liver Disease.(*15*) In BA, there have been reports of epithelial signatures consistent with both transdifferentiation but also LGR5+ cell subsets.(*8, 16*) However, the exact role, distribution and functional relevance of these epithelial states to the disease severity and outcomes, remains unclear.

Among the many cell types in the BA ‘ecosystem’, the role of the intrahepatic lymphatic vasculature remains poorly understood. The common referral of the ‘portal triad’ instead of tetrad underscores the negligence to acknowledge their relevance, despite the liver being among the largest producers of lymph generating 25-50% of lymph that passes through the thoracic duct back into the body’s circulation.(*17*) Outside the liver, the lymphatic vasculature is well established as a regulator of fluid homeostasis, wound healing, and immune surveillance, with CCL21 recognized as a key mediator of lymphatic immune cell trafficking.(*17–19*) A reactive lymphangiogenesis has been proposed in chronic liver diseases.(*20*) Beyond merely responding to injury, here, we assess an active role of lymphatic endothelial cells (LEC) at the time of KPE to modulate the future ability of intrahepatic bile ducts to drain bile and evade transplantation.

Here, we provide a single-cell, spatial, and 3D-imaging-based atlas of early BA livers retrospectively stratified for outcome after KPE. Unbiased analysis identifies hepatic lymphatic dysplasia as an early determinant of rapid disease progression towards transplantation in BA. Functional validation in tissue- and iPSC-derived organoids and assembloids reveals that excessive lymphatic CCL21 signals on inflammation-primed liver epithelium and shapes its fate in BA.

## RESULTS

### Multiscale profiling of BA identifies early determinants of divergent outcomes

To identify molecular, tissue and functional determinants of intrahepatic BA outcome, we sampled liver tissue at three German university hospitals at the timepoint of KPE and stratified for clinical outcome thereafter. We compared BA and non-BA cholestatic control patients that include Choledochal Cyst, Alagille Syndrome, and Progressive Familial Intrahepatic Cholestasis using a variety of experimental modalities to dissect BA-specific phenomena from general cholestatic effects. To link cell-specific molecular programs to multicellular tissue circuits, we combined single-nucleus RNA-sequencing (snRNA-seq) analysis of cryopreserved samples with multiplexed (mpIF) and 3D-immunofluorescence, spatial Stereo-sequencing (Stereo-seq) and a variety of *ex vivo* profiling and mechanistic organoid studies (Fig. 1A-B). To delineate biological processes governing epithelial outcome after KPE, we retrospectively stratified for long native liver survival vs. infants with rapid progression toward liver transplantation (LTX) in the first year due to biliary epithelial dysfunction, independent of viral infections or other possibly confounding factors. We classified patients achieving jaundice clearance, defined as normalization of serum bilirubin levels by 100 days post-KPE, and sustained native liver survival as a ‘mild BA’ phenotype (n=14). Infants with persistent hyperbilirubinemia at 100 days post-KPE and LTX within their first year of life were classified as ‘severe BA’ patients (n=12). We deemed biosampling at KPE as ‘pre-outcome’ and used those samples for snRNA-seq, Stereo-seq, mpIF and tissue-cleared 3D-light sheet microscopy (Fig.1B). For functional validation, we performed ‘post-outcome’ biosampling from BA and non-BA liver explant tissue received during LTX. From these tissues, we generated intrahepatic cholangiocyte organoids (ICO), isolated lymphatic endothelial cells (LECs) and conditioned media from minced tissue for proteomic analysis of soluble disease tissue mediators. For functional validations requiring multiple cell types – i.e. cholangiocytes and hepatocyte – we also used iPSC-derived fetal liver-like organoids (FLO)(*21*) and hepatocyte-like cells (iHeps)(*22*). Notably, patient age at KPE, histopathological fibrosis levels (as measured by the ISHAK score) and direct bilirubin levels did not significantly differ between mild and severe BA patients (Fig.1C), enabling the identification of jaundice clearing mechanisms specifically. Mean age at LTX was 6.6 (±2.2 SD) months in the severe BA group. Mean duration of native liver survival in the mild BA group is 6.8 (±2.7 SD) years, with only one patient being transplanted during the ongoing study. In sum, this study design offers an outcome-stratified BA cohort, as the basis for the different analyses that follow.

**Fig. 1.**
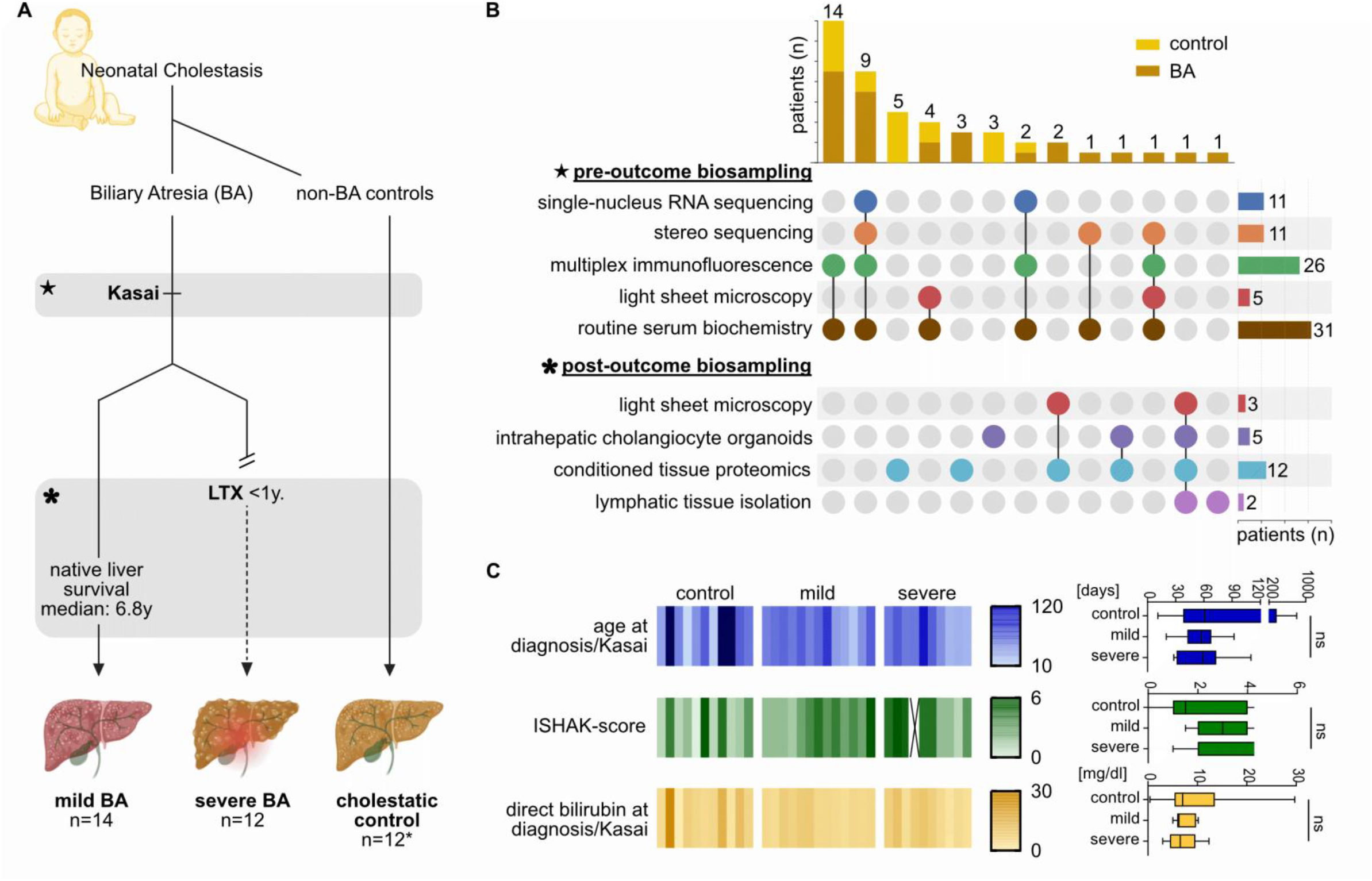
Multiscale profiling of outcome-stratified biliary atresia to identify early disease drivers undetected by routine diagnostics. **A:** Clinical stratification of patients with neonatal cholestasis. *An additional eight non-cholestatic patients (AIH, metabolic disorder, portal veine thrombosis, adult liver tumor) were sampled at post-outcome sampling for generation of intrahepatic cholangiocyte organoids (ICO) and conditioned media. **B:** UpSet-Plot shows the number of patients sampled for one or multiple experimental techniques, respectively, at diagnosis/Kasai (pre-outcome biosampling) and LTX (post-outcome biosampling). **C:** Clinical characteristics and serum biochemistry of patients sampled at pre-outcome sampling shown in a heatmap and summarized as minimum to maximum with interquartile range in boxplots. One-way ANOVA, ns = not significant.

### Maladaptive lymphatic dysplasia is an early determinant of poor outcome after KPE

To dissect the heterogenous cell types and states determining clinical outcomes, we leveraged snRNA-seq from a multi-center and retrospectively outcome-stratified patient cohort. Using mechanical homogenization, avoiding enzymatic dissociation, which typically biases against matrix-embedded and fragile stromal, endothelial and epithelial cell types in fibrotic tissue, we identified that most cell types were conserved across the three groups, including rarer cell types, e.g. lymphatic endothelial cells, T and B lymphocytes (Fig.2A, Fig.S1A-F for annotations).

**Fig. 2.**
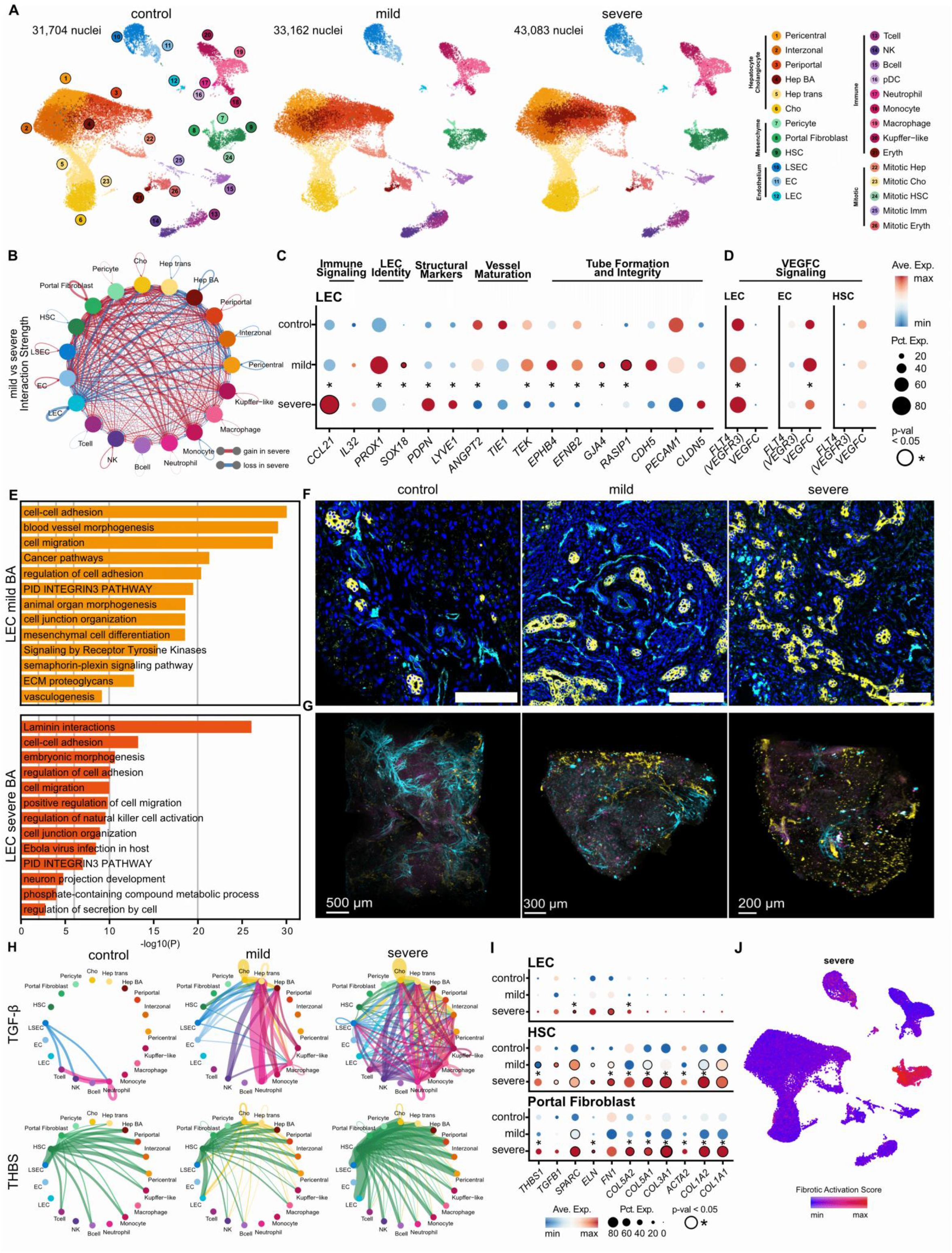
CCL21-high, dysplastic lymphatic endothelial phenotype marks early BA with poor outcome. **A:** Uniform manifold approximation and projection (UMAPs) show cell clusters in the different patient groups. **B:** Chord diagram shows differential interaction strength between mild and severe BA. **C:** Top differentially expressed genes of LECs. **D:** VEGFC signaling in BA. For dot plots: colour-coded expression levels, size-coded percentage of cells expressing each gene. **E:** Top enriched pathways based on interaction mediators in mild and severe BA LECs. **F:** Representative mpIF images of ductular cells (CK19, yellow) and LECs (PDPN, cyan blue) in portal areas of control, mild BA and severe BA livers. **G:** Representative images of light sheet microscopy of whole-mount IF stainings show LECs (PDPN, cyan blue), ductular cells (CK19, yellow) and leukocytes (CD45, magenta). See supp. material for videos. **H:** Chord diagram of *TGFβ* and *THBS* interactions across all cell clusters. **I:** Contribution of LECs, HSCs and portal fibroblasts to fibrotic signaling. **J:** Activation score of the fibrotic signaling genes in Fig2I in the severe case. **C-J:** Significant differences are depicted by black circles if a condition is significantly regulated compared with controls and an asterisk if a significant difference is noted between mild and severe BA.

While the overall inferred cell-cell interactions escalated with disease severity (Fig.S2A-C), LECs lost most of their physiological interactions in severe BA when compared to mild BA (Fig.2B). A detailed assessment of expression modules for LEC identity, function and structure revealed a loss of *PROX1, SOX18, ANGPT2, EPHB4 and EFNB2* in severe BA (Fig.2C). Surprisingly, severe BA LECs mount a selective surge in *CCL21* and *PDPN*. Mild BA LECs displayed an opposite pattern, indicating critical differences of lymphatic endothelial identity, vasculogenesis, remodeling, and integrity across BA outcomes.(*23, 24*) Interestingly, VEGF-C, the main stimulus of lymphangiogenesis was significantly decreased in endothelial cells (EC) in severe BA, while VEGF-C-receptor expression (*FLT4*) was significantly increased in LECs – potentially reflecting a failed compensatory response (Fig. 2D). When comparing most enriched biological processes between mild and severe BA LECs that were derived from the cell-cell interaction members, the mild BA LEC top terms ‘blood vessel morphogenesis’ and ‘vasculogenesis’ were missing in severe BA LECs, which suggests insufficient lymphangiogenesis in line with the aberrant gene signature (Fig.2E). The top gene ontology (GO) term of severe BA LECs – ‘laminin interactions’ – may further suggest their contribution to ECM remodeling.

In accordance with the unstable *PDPN*^high^, *CCL21*^high^ maladaptive LEC state in severe BA, we validated high PDPN+ and continuous CCL21-staining patterns in mpIF stainings (Fig.S3A-D). Further, mpIF of 28 FFPE liver sections confirmed insufficient formation of lymphatic endothelial vasculature. While mild BA livers show a consistent pattern of expanded lamellar-like lymphatic endothelial structures intercalated with the CK19+ epithelium as a sign of portal lymphangiogenesis, severe BA livers were characterized by an increased number of malformed, clumped PDPN+ aggregates in the periportal area and reduced numbers of elongated, endothelial structures (Fig.2F). Whole mount IF and light sheet microscopy confirmed a sparse and fragmented lymphatic network most pronounced in severe BA, consistent with a dysplasia phenotype (Fig.2G). In comparison, livers of cholestatic-fibrotic controls showed a dense, organized and highly branched lymphatic network.

A dysplastic, CCL21^high^ lymphatic vasculature suggested altered immune cell trafficking in severe BA. MpIF indicated consistent accumulation of CD3+ lymphocytes in BA livers and specifically at close distance to LECs. However, differences were not statistically significant compared to mild BA patients (Fig.S3D). After observing lymphatic dysplasia in severe BA, mpIF further confirmed that severe BA patients mount ductular reactions early on. Consistently, 7 out of 9 severe BA-patients but only 1 out of 10 mild BA patients classified as CK19^high^ tissue indicating a significant surge in CK19+ ductular cells in severe BA, when compared to mild BA patients with better clinical outcomes (Fig.S3C+D). The observations of largely co-localized lymphatic and biliary dysplasia in the expanding portal areas of severe BA patients suggested that interactions between CCL21^high^ lymphatic vasculature and cholangiocytes may be possible.

Further differential pathway inference showed thrombospondin (*THBS*) and transforming growth factor beta (*TGFβ*) enriched in severe BA (Fig.2H). Surprisingly, both *THBS* and *TGFβ,* previously described as important regulators of liver injury and fibrosis,(*25,26*) were amongst the few upregulated genes in LECs from severe BA indicating a fibrogenic LEC state in those patients proceeding towards poor clinical outcome (Fig.2H-J). While the inferred number of cell populations involving hepatic stellate cells (HSCs) and portal fibroblasts were larger, this data, together with LEC pathway enrichment (Fig.2E) discussed earlier, shows that LECs contribute to local fibrogenic circuits, which is relevant given their portal location in proximity to cholangiocytes.

### SnRNA-seq identifies epithelial cell states associated with early LTX in BA

Across clinical strata, snRNA-seq identified highly plastic epithelial cell states (Fig. 2A; Fig.3A-G; Fig.S1A-F). We identified hepatocyte-to-biliary spectrum cell states (Hep trans), as a broad adaptive response in all cholestatic injuries (Fig. 3A). However, in severe BA livers the Hep trans state pivoted the strongest towards biliary transdifferentiation states (i.e. *SOX9*/*HNF1B*^high^, *HNF4A*^low^) and involved LGR5. This finding demonstrated that transdifferentiation is found across cholestatic injuries but that severe cholestatic injury exacerbated the liver’s plasticity response and engages the Wnt/β-catenin-responsive progenitor activity (Fig.3D-F). Trajectory analysis inferred that periportal and interzonal hepatocytes may participate in transdifferentiation programs (Fig. 3B). Notably, we also found another distinct BA-specific hepatocyte subset enriched in severe BA. This ‘Hep BA’ cell state was characterized by the instability of hepatocyte identity (*HNF4A*^low^) and transcriptional programs high in acute-phase, metal-/redox-stress and ECM-crosslinking signatures, e.g. *CRP, SAA1, LOXL1* and *TGM2* (Fig.3C-E; Fig.S1D). These Hep BA cells were also enriched for GO terms like ‘wound healing’, ‘viral response’, ‘innate immune response’ (Fig.S4A). We then sought to check whether all other hepatocytes could glean the Hep BA phenotype by calculating a score for the top Hep BA genes (Fig.S4B). Interestingly, we found a stepwise increase of cells taking on this phenotype with increasing disease severity.

**Fig. 3.**
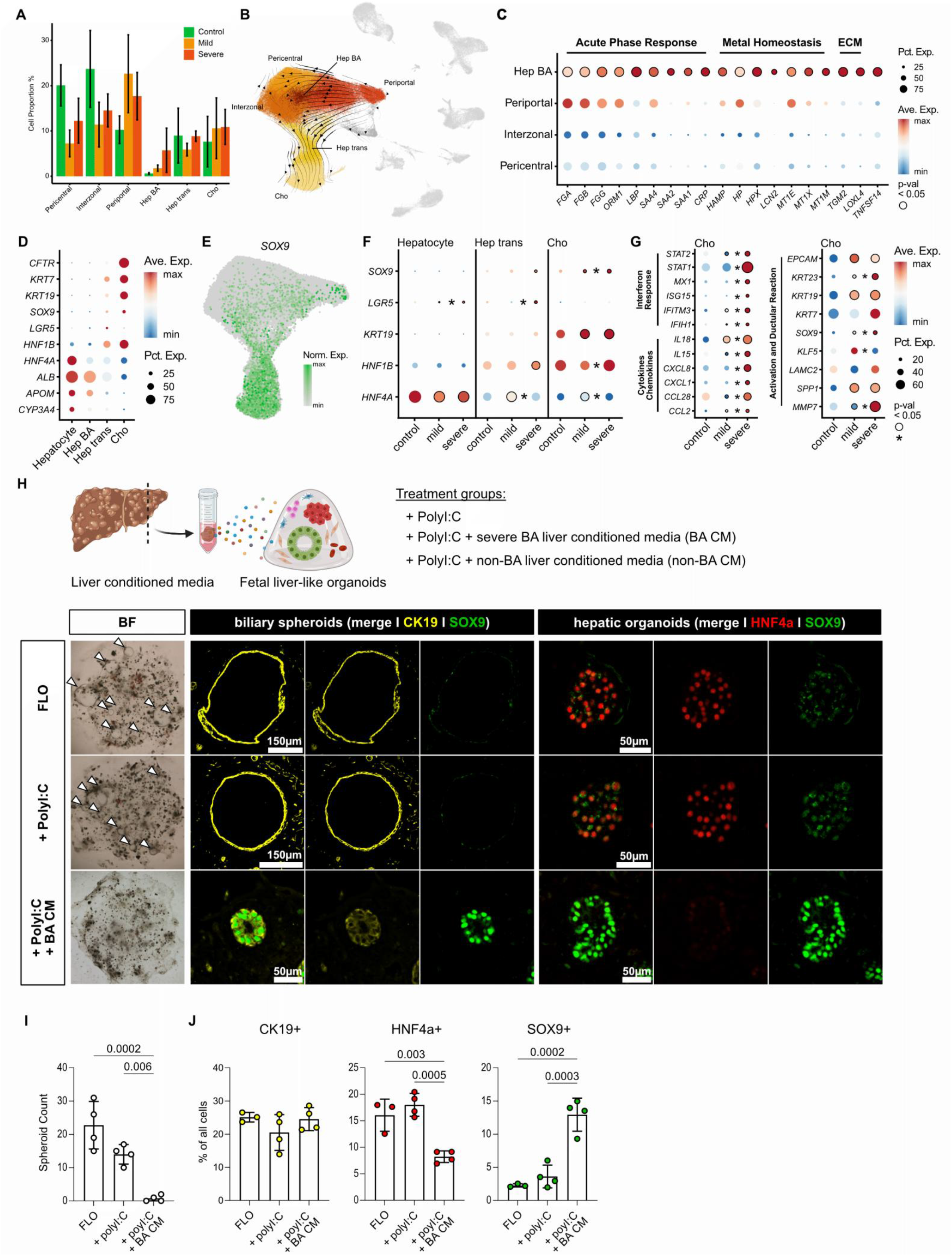
Epithelial plasticity and immune-reactive cholangiocyte reprogramming in BA and developmental organoid models of BA. **A:** Cellular proportions across disease severities depicted as mean ± SEM. **B:** Pseudotime trajectory projected onto the UMAP with arrows indicating the inferred differentiation paths. **C:** Top differentially expressed genes of Hep BA. **D:** Lineage marker expression across hepatocytes and cholangiocytes. **E:** *SOX9* expression in hepatocytes and cholangioyctes. **F:** Lineage marker expression in hepatocytes and cholangiocytes split by disease severity. **G:** Top differentially expressed genes of cholangiocytes. **H:** Experimental set-up of iPSC-derived fetal liver-like organoids (FLO) treated with polyI:C +/-conditioned media (CM) of BA or non-BA liver explants. Bright Field (BF) and immunofluorescence (IF) images of FLOs under respective treatments. **I:** Bar graphs show mean±SD for number of spheroids per dome. **J:** Bar graphs show mean±SD for % of CK19+, HNF4a+ and SOX9+ cells respectively. **A-G:** Significant differences are depicted by black circle outlines if a condition is significantly regulated compared with controls and an asterisk if a significant difference is noted between mild and severe BA. For dot plots: colour-coded expression levels, size-coded percentage of cells expressing each gene. **I+J:** One-way ANOVA results are only indicated if p<0.05.

Importantly, regarding the cholangiocyte response across clinical BA strata, we identified a unique state enriched in severe BA patients characterised by a broad Interferon-responsive and chemotactic program suggestive of a lymphoid immune response and possibly post viral state (Fig.3G, Fig.S4D). Condition-specific GO terms confirmed further differences with ‘wound healing’, ‘cell-substrate adhesion’, and ‘response to TGF-β‘, being shared by mild and severe cholangiocytes, while ‘response to virus’, ‘chemotaxis’, and ‘tissue remodeling’ were specifically enriched in severe BA. Interestingly, these severe BA cholangiocytes were again highly expressing *SOX9* as well as the BA-specific epithelial gene product and diagnostic biomarker *MMP7* (*4*) (Fig.3G, Fig.S4D). Of note, MMP7 expression levels of cholangiocytes derived from mild BA livers were equal to those from cholestatic controls. Because MMP7 is, thus far, not well established as severity marker after KPE, our results add this new dimension to it. While cholangiocytes from severe BA exhibited a highly activated state, they showed a significant loss of KLF5, a transcription factor required for biliary epithelial remodelling and integrity in cholestatic liver injury.(*27*)

Aside from the epithelial cells, we also sought to characterize the disease severity differences in key mesenchymal and myeloid cell types of the BA liver (Fig.S4D). HSCs exemplified a shared ‘wound healing’, ‘collagen organization’ and ‘viral response’ phenotype in BA regardless of severity. Meanwhile in severe BA, we find ‘interferon response’, ‘TOR signaling’, and ‘smooth muscle cell migration’ that may indicate mesenchymal cell activation. For Kupffer-like cells, the BA condition showed strong enrichment of terms involved in the immune response, viral process, and cell adhesion. In severe BA, we find ‘leukocyte proliferation’ and ‘response to dsRNA’ terms to be uniquely enriched (Fig.S4D). Across all cell types, IFN signaling indicative of a lymphoid immune response was enriched in severe BA compared to mild and control (Fig. S4E).

### Fetal liver-like organoids recapitulate SOX9^high^ structural atresia and HNF4A loss

BA has a presumed fetal origin, yet no fetal model has captured how hepatocytes and cholangiocytes jointly respond to a BA-specific microenvironment. We established a BA-specific multi-compartment fetal liver-like organoid (FLO) system, combined with patient-derived liver secretome, to test what injury is sufficient to drive the epithelial plasticity and biliary lumen collapse (‘atresia’) seen in patients (Fig.3H). After detecting an IFN-high signature in severe BA, we used the synthetic dsRNA analogue polyinosinic:polycytidylic acid (polyI:C) to model the convergent, virus-independent innate response, also previously shown to induce the BA phenotype.(*28*) We combined this injury with the addition of conditioned medium (CM) derived from minced BA and non-BA liver tissue, sampled at LTX (post-outcome biosampling) (Fig.3H). Intriguingly, different from the literature above, polyI:C treatment alone, had no significant effect on lumen-forming biliary phenotypes or epithelial plasticity in FLOs (Fig.3H). Yet, additional treatment of differentiated multilineage FLOs with CM from severe BA patients (but not non-BA controls, Fig.S5A+B), resulted in a complete biliary lumen-collapse (‘atresia’) (Fig.3I). Both BA and non-BA CM treatment led to the transdifferentiation of HNF4A+ grape-like hepatocyte clusters to SOX9+ cells (Fig.3J; Fig.S5A+C). At the same time, only the addition of BA CM largely suppressed HNF4A in hepatic organoid assemblies, when assessed at protein level using mpIF on FLOs (Fig.3J; Fig.S5C). Thus, a BA-like model that combines a developmental, multicellular organoid with patient tissue secretome provides evidence that BA-specific niche signaling establishes cellular cues that promote disease and not innate epithelial programs alone drive BA-associated cell states.

### Spatial Stereo-sequencing offers insights into adaptive and maladaptive lymphatic cell neighborhoods

Because snRNA-seq dissociates tissue architecture, we applied Stereo-seq to intact liver sections from cholestatic control and BA patients to spatially resolve the portal lymphatic niche at single-cell resolution. Stereo-seq captured the cellular complexity of the liver in both cholestatic control and BA patients (Fig.4A+B, Fig.S6A-D) by detecting 434,563 cells and 15,679 genes and resolving hepatocyte-dominated hepatic regions and portal fibroblast enriched portal regions (Fig.S6E). Cholangiocytes formed duct-like structures surrounded by stromal, endothelial, and immune cells, while portal regions became expanded and interconnected in BA, all consistent with general understanding of BA, validating our spatial data. Cellular community analysis identified seven communities across tissues, two of which were dominated by cholangiocytes (Fig.4C+D, Fig.S7A). Community 5 comprised mainly cholangiocytes and Hep trans cells, suggesting that transitional cells may acquire biliary identity. GO terms were enriched for ‘cell polarity’, ‘movement’, ‘cell-cell junctions’, ‘morphogenesis’, and ‘development’. Community 3 comprised portal area associated populations, including portal fibroblasts, endothelial and lymphatic endothelial cells, and immune cells, and was enriched for ‘ECM assembly’, ‘vessel morphogenesis’, ‘TGF-β production’, ‘chemotaxis’, and ‘development’ (Fig.S7B).

**Fig. 4.**
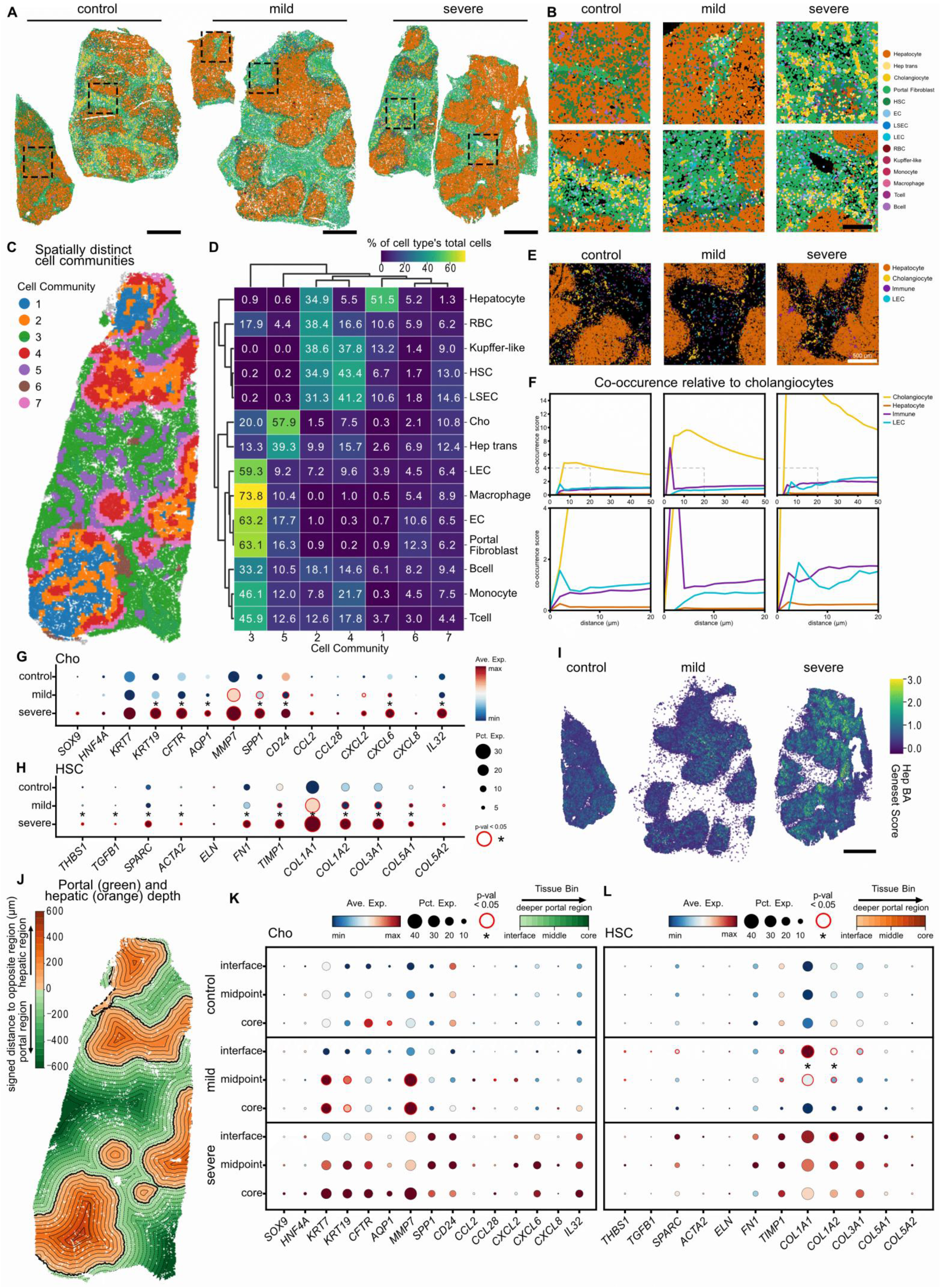
Spatial in situ-sequencing identifies early cell communities associated with poor outcome. **A:** Spatial distribution of identified cell types. Scale bar = 1000 µm. **B:** Expanded view of bounded box areas. Scale bar = 150 µm. **C:** Spatial distribution of identified cellular communities in a severe BA sample. **D:** Composition of cellular communities across all samples. **E:** Expanded views of portal areas in selected samples. **F:** Cellular co-occurrence analysis relative to cholangiocytes. **G:** Gene expression differences across severity in cholangiocytes and **H:** HSCs. Red circle indicates significant difference compared to controls. Asterisk indicates significant difference between mild and severe BA. **I:** Spatial distribution of Hep BA geneset score expression. **J:** Signed distance measurement of depth relative to hepatic and portal regions. Concentric rings represent 100µm bins. Maximum depth is 600µm from the interface (thick lines). **K:** Expression of ductular reaction and inflammatory markers in cholangiocytes when binning the regional depth into 3: interface, midpoint and core. **L:** Expression of profibrogenic markers in HSCs when binning the regional depth. **K+L:** Red circle indicates significant difference between the least activated region vs. the midpoint and most activated region. Asterisk indicates significant difference between the midpoint and most activated region.

Cellular co-occurrence analysis revealed greater co-occurrence of cholangiocytes with other cholangiocytes with increasing disease severity, consistent with our previous findings of expansion during ductular reaction of severe BA (Fig.4E+F, Fig.S7C). Co-occurrence with lymphatic endothelium also appeared increased in severe BA but was not a consistent phenomenon across all samples. These patterns may partly reflect changes in cell abundance. Spatial profiling confirmed that inflammatory mediators and ductular reaction, including SOX9 and MMP7, were most strongly expressed in severe BA cholangiocytes (Fig. 4G). HSCs exhibited the highest expression of fibrogenic genes in severe BA (Fig.4H), collectively corroborating the snRNA-seq findings at a spatial level.

Although data sparsity precluded identification of the Hep BA subcluster in the Stereo-seq dataset, projection of its top differentially expressed genes revealed a progressive increase in the Hep BA score with disease severity, validating the trend observed by snRNA-seq (Fig.4I, Fig.S7D). An interferon-response gene score similarly increased in severe BA, spatially validating the severity-associated interferon signature (Fig.S7E).

Spatial analysis along the hepatic–portal axis showed that established hepatocyte zonation markers, including *CYP3A4*, *GLUL*, and *HAL*, followed their expected expression patterns. The cellular communities localized predominantly to hepatic or portal regions, or at their interface (Fig.4J, Fig.S7F-H). To resolve regional differences in cell states, we delineated the hepatic and portal regions and calculated signed distances from their interface, using 100 μm bins radiating along the hepatic–portal axis (Fig.4J, Fig.S7F). Cells were then classified within their respective regional niches as interface, midpoint, or core (Fig.4K+L). This spatial stratification largely recapitulated the cholangiocyte signatures shown in Fig.4G, while revealing additional regional heterogeneity. In severe BA, interface cholangiocytes expressed high levels of *SPP1* and *CD24*, whereas midpoint-region cholangiocytes showed increased ductular reaction and inflammatory mediators, including *CXCL2*, *CXCL6*, and *CXCL8* (Fig.4K), consistent with our snRNA-seq data (Fig.3G). Core-region cholangiocytes showed the strongest expression of ductular reaction markers, including *KRT7*, *KRT19*, and *MMP7*. Hep trans cells exhibited a similar spatial pattern, consistent with acquisition of a stronger biliary identity (Fig.S7I). Finally, HSCs showed greater profibrogenic activity at the hepatic–portal interface (Fig.4L), suggesting that the local microenvironment contributes to HSC activation and architectural remodeling in BA liver tissue.

### Lymphatic endothelium controls SOX9- and MMP7-states of immune-primed cholangiocytes

Post-outcome biosampling obtained tissue and cell isolates from BA and non-BA liver explants for functional validation using ICOs and ICO-LEC-assembled organoids. For in-depth profiling of the soluble BA microenvironment, CM was obtained as mentioned above and profiled with Proteomics, LegendPlex^TM^ and ELISA.

Proteomics detected a total of 1823 proteins of which 134 were unique to BA (Fig.S8A-C). Overall pathway analysis enriched ‘epithelial cell development’, ‘immune system’, ‘ECM proteoglycans’ and other pathways in BA compared to non-BA. Lipid metabolic processes and lysosome activity were also enriched, reflecting a high bile acid metabolism. When further comparing CM from mild (late LTX) and severe (early LTX) BA patients, the latter enriched pathways concerning innate immune system, ECM receptor interaction, fibrillar collagen trimer and cadherin binding as well as lysosome. ICAM-1 was more than 10-fold higher in severe BA CM than in mild, and CD44 was uniquely detected in severe BA, possibly resembling inflammatory endothelial activation and immune cell enrichment. Interleukin (IL)-18 was detected exclusively in CM from severe BA, but not in CM from mild BA, echoing reports linking IL-18 to post-Kasai outcomes.(*29*) LegendPlex^TM^ detected no significant differences between BA and non-BA secretome for TNF-alpha, IL-1B, IL-17A, and IL-8 surprisingly (Fig.S8D). Interferon-Gamma (IFN-γ), Interferon-alpha2 (IFN-α2), IL-12-p70, IL-33, IL-6 and MCP-1 were significantly higher in BA.

Given the previous observation of extensive biliary and lymphatic dysplasia in the portal areas of severe BA patients and the epithelial plasticity observed in BA-secretome-treated FLOs, we hypothesized that LECs from severe BA may directly affect the biliary epithelium and induce epithelial state changes *via* CCL21. To model lymphatic-to-epithelial effects, we isolated LECs from BA liver explants, leveraging magnetic bead-based fractionation, assembled them with non-BA ICOs in a 3D assembloid and observed *SOX9*^high^ *LGR5*^high^ biliary progenitor state, as in the severe BA tissue response highlighted earlier (Fig.5A, Fig.3D-G). Direct treatment of ICOs with recombinant CCL21 alone partially recapitulated the SOX9^high^ epithelial state and confirmed that cholangiocytes can respond to CCL21 (Fig.S9A+B). As not all patient-derived ICO lines were responsive to CCL21, we found that the permissive epithelial state for CCL21 was determined by the condition from which the ICOs were derived. Specifically, CCL21-sensing ICOs were derived from livers with histopathological evidence of immune cell infiltration and ductular reaction (Fig.S9B).

**Fig. 5.**
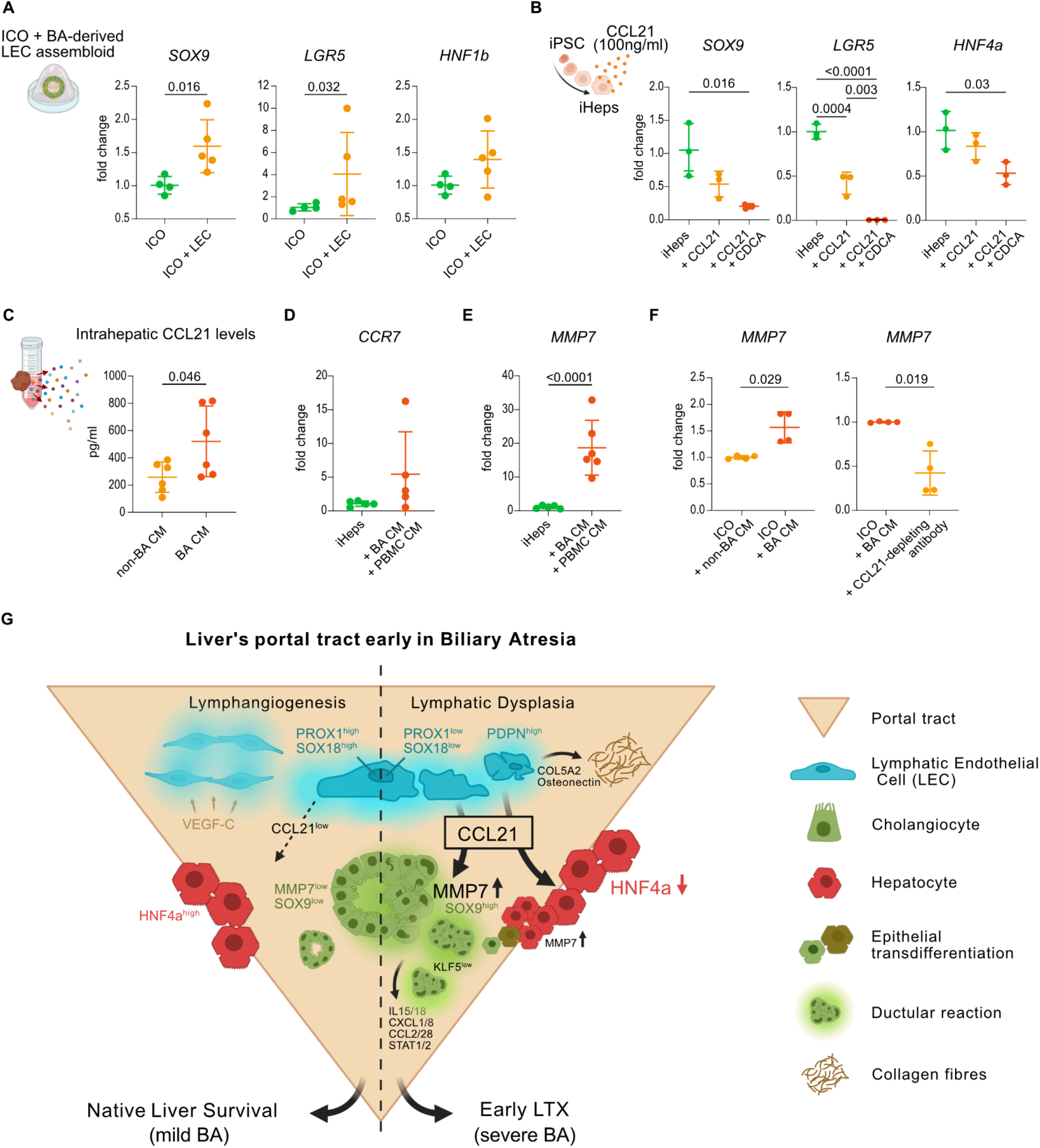
CCL21 shapes MMP7+ epithelial states *ex vivo*. **A:** Nine day co-culture of ICOs derived from non-BA patients (n=2) with LECs isolated from BA explant livers (n=2). Graphs show mean±SD of fold change up-/downregulation of respective genes in ICOs. **B:** CCL21 treatment (100ng/ml) of iPSC-derived hepatocytes with and without addition of 200µM CDCA. Graphs show mean±SD of fold change up-/downregulation of respective genes. **C:** CCL21 ELISA of CM from non-BA (n=6) and BA (n=6) liver explants measured liver-derived CCL21. **D:** Treatment of iPSC-derived hepatocytes with BA CM and pI:C-primed PBMC CM. Graph shows mean±SD of fold change up-/downregulation of CCR7. **E:** Treatment of ICOs with non-BA and BA CM. Graphs show mean±SD of fold change up-/downregulation of MMP7. **F:** Treatment of ICOs with non-BA and BA CM +/-CCL21-antibody. Graphs show mean±SD of fold change up-/downregulation of MMP7. **G:** Graphical summary of the proposed mechanisms underlying mild versus severe BA. **A-F:** One-way ANOVA results are only indicated if p<0.05.

Following up on the hepatocyte states identified in severe BA – transdifferentiating (Hep trans) and identity-destabilized (Hep BA) hepatocytes (Fig.3A–D) – we treated iPSC-derived hepatocytes with CCL21 and interestingly observed alterations of their transcriptional profile, indicating CCL21-sensing in hepatocytes. Suppression of *SOX9, LGR5* and *HNF4A* indicated broad collapse of both biliary and hepatocyte identity programs rather than a directed lineage transition (Fig.5B). While bile acids have been shown to regulate cholangiocyte plasticity and promote biliary differentiation of liver progenitor cells, their potential to alter hepatocyte identity in a cholestatic milieu remains unclear.(*30, 31*) As bile acids largely determine the hepatocyte microenvironment in BA, we combined the CCL21 treatment with Chenodeoxycholic acid (CDCA) at a non-cytotoxic concentration and observed exacerbated identity loss relative to CCL21 alone.

Intrahepatic CCL21 was elevated in BA liver secretome (Fig.5C; Fig.S9C), consistent with the CCL21^high^ LEC transcriptome and mpIF staining observed at the earlier pre-outcome timepoint in severe BA patients, indicating sustained CCL21 elevation along disease progression. To model the effects of disrupted lymphatic signaling on the liver epithelium within the complex cholestatic and inflammatory BA environment ex vivo, we treated hepatocytes with CCL21^high^ BA CM in combination with indirect co-culture with polyI:C-primed PBMCs (Fig.5D+E; Fig.S9D; pIC-PBMC CM). This BA-like environment induced upregulation of C-C chemokine receptor type 7 (*CCR7*) in some but not all patient epithelia. Further, it led to a surge of the BA-specific *MMP7* (Fig.5E). Assessing the translational value of CCL21-modulating strategies to improve epithelial outcomes, CCL21-neutralizing antibodies normalized the BA-associated *MMP7^-high^*-state in ICOs (Fig.5F; Fig.S9E), suggesting that CCL21 is not only a marker of the dysplastic lymphatic niche but also a tractable effector of BA epithelial states (Fig.5G).

## DISCUSSION

BA follows a multifactorial pathophysiology and is the leading cause of pediatric liver transplantation worldwide.(*1, 32*) To dissect the molecular and structural tissue features that bias patient outcomes of BA early on, we profiled infant liver tissue obtained at KPE that was retrospectively outcome stratified. Thereby, we distinguished distinct intrahepatic BA phenotypes and identified early intrahepatic determinants of rapid progression towards liver transplantation in BA.

Here, we establish across a series of experimental modalities (spanning transcriptional, mpIF, liver tissue secretome-based ELISA, functional experiments in organoids) that early CCL21^high^ lymphatic endothelial dysplasia is a hallmark of BA patients who proceed with poor outcome. Structurally, severe BA exhibited a maladaptive aggregation of lymphatic cells and a loss of lymphatic endothelial morphology continuity, whereas mild BA, although also affected relative to cholestatic controls, displayed an adaptive lymphangiogenic response.

Interestingly, we found that lymphatic endothelium signals on liver epithelium *via* CCL21 and induces a SOX9^high^ LGR5^high^ biliary progenitor response and most interestingly an MMP7^high^ epithelial state in cholangiocytes. This is relevant since a landmark study and subsequent validations established MMP7 elevation not only as a BA-specific phenomenon and biomarker, both at serum and hepatic transcript level, but also as mechanistically relevant in BA mouse models.(*2, 4, 9, 10, 33*) Yet here we show an upstream lymphatic-to-epithelial circuit, based on findings that soluble mediators as CCL21 can shape biliary epithelial responses, contributing to maladaptive remodeling or tissue repair.(*34*) We demonstrate that hepatic lymphatic tissue can exert these ectopic epithelial effects after prior BA-specific inflammatory licensing. *Ex vivo*, we suggest that a CCL21 receptor, CCR7, can potentially be induced on epithelial cells exposed to the tissue supernatant of severe BA livers. Yet, CCR7 expression levels vary and may be controlled by epigenetically determined permissive states. Beyond CCL21’s role in homing immune and trafficking metastatic cells toward the lymphatic system,(*35*) our data lends well into reports of ectopic CCL21 sensing in liver fibrosis (on myofibroblasts).(*36*)

CCL21-signaling beyond the lymphatic endothelium is dependent on glycosaminoglycan binding in the ECM,(*19*) thus CCL21-accessability should be considered in subsequent studies. Interestingly, cholestasis-associated increase in proteoglycans on cholangiocytes and HSCs may alter the local retention and spatial presentation of LEC-derived CCL21, thereby modulating epithelial exposure to this chemokine.(*37*) Consistent with this finding, ‘ECM proteoglycans’ pathway was enriched in BA liver’s secretome relative to non-BA controls.

Our findings of maladaptive lymphatic dysplasia in severe BA may offer a shared mechanism for disease progression in severe inflammatory cholestasis, given a preprint report on lymphatic changes in PSC, although our focus on CCL21-effects on immune licensed-epithelium has a different angle.(*38*) Different from the report on lymphatic endothelium after bile duct ligation in mice, our mpIF analysis did not reveal alterations of immune cell trafficking or accumulation around dysplastic lymphatic vasculature. Of note, we were comparing BA patients to cholestatic control patients and not to healthy livers and differences in immune cell accumulation around LECs were not significant. The report further linked the increase of TGF*β* in LECs to reduced LEC density and drainage capacity.(*38*) Interestingly, in our outcome-stratified BA cohort, enrichment of TGF*β*, a driver of epithelial plasticity, was not significant in severe BA LECs. However, we observed partial fibrogenic activity in LECs of severe BA patients with enrichment of SPARC (Osteonectin) and COL5A2. Thus, LECs may contribute to liver fibrosis in BA. (*14, 39*) In HSCs, TGFβ was also not differentially expressed between HSCs from mild and severe BA, supporting our foundational observation in Fig. 1, that fibrosis is not an obvious driver of outcome. Overall, our study suggests that the lymphatic compartment in BA is not only a bystander injury but actively engages with the epithelium with partial contribution to fibrogenicity.

Consistent with the central role of the liver epithelium in BA pathophysiology, we observed highly plastic epithelial cell states with distinct phenotypes characterizing severe BA. SnRNA-seq detected hepatic-to-biliary transdifferentiation unspecifically in BA and cholestatic controls, yet the epithelial spectrum is more nuanced in BA with increased SOX9/HNF1b expression, which was most prominent in transdifferentiating hepatocytes in severe BA.(*14*) Interestingly, severe BA also engaged LGR5+, distinct from the direct transdifferentiation reported in adults.(*15*) Functionally, we provide evidence for LECs stimulating epithelial progenitor reprogramming, engaging LGR5, using a novel assembloid system. This offers a new possibility of a lymphatic rheostat of the liver’s LGR5+ progenitor subset, previously reported in cholestatic liver injury.(*13*) Our FLO *in vitro* experiments recapitulated epithelial plasticity towards SOX9^high^ states under severe BA-‘secretomes’ and revealed that this state is not driven by intrinsic epithelial programs or viral mimetic-polyI:C alone. In addition, FLOs, exhibited biliary lumen-collapse despite increased SOX9 expression, similar to the ductular reaction in BA, and HNF4a loss of the hepatic compartment.(*40*) Consistently, severe BA enriched a distinct hepatocyte dedifferentiation and immunogenic state exhibiting HNF4A^low^ identity loss, acute reactive and inflammatory programs (Hep BA) as identified by snRNA-seq. Interestingly, CCL21 treatment of hepatocytes in a cholestatic milieu *ex vivo* induced this Hep BA-like state, that included a loss of *HNF4A*, *SOX9* and *LGR5*. A recent transcriptional study on BA also linked poor native liver survival to epithelial plasticity, however molecular analysis was done at late stage on explant livers and could not recapitulate diverging outcomes at the timepoint of KPE.(*41*)

Lastly, we detected a unique cholangiocyte signature in severe BA that was characterized by proinflammatory and profibrotic programs with extensive expression of chemokines and cytokines, highlighting their active role in an inflammatory hub in BA pathophysiology. Additionally, MMP7 was significantly increased in severe compared to mild BA cholangiocytes at the timepoint of KPE.

One limitation of our findings is that the exact inflammatory priming of CCL21-sensing may involve factors beyond CCL21, which requires further analyses, but could involve additional cytokines, chemokines or extracellular vesicles. Regarding functional experimentation, the standard rhesus rotavirus (RRV)–induced mouse model of BA is restricted to a single mouse strain, and species-specific differences in the CCL21 system, including a third mouse Ccl21 gene (Ccl21c) with no human ortholog and predominantly non-lymphoid expression, may limit its translational relevance for validating CCL21-dependent mechanisms in BA. Moreover, biliary injury is induced postnatally in the RRV mouse model, whereas mounting data suggests fetal onset in humans,(*5, 42, 43*) further limiting its translational relevance. Therefore, we pursued a human tissue-centered approach, combining patient-derived tissue analyses with human liver- and iPSC-derived organoid models for ex vivo functional validation, thereby enabling the investigation of human BA pathophysiology in the most relevant context.

In summary, our multiscale atlas indicates that outcome after KPE is shaped by intrahepatic cell states already present at diagnosis raising the possibility that distinct pathogenic mechanisms may underlie the clinical diagnosis of BA. Our findings specifically underscore the previously underappreciated role of the lymphatic niche within the ‘portal tetrad’ in BA progression, offering potential opportunities for prognostic stratification and therapeutic interventions which should be addressed in future prospective studies.

## MATERIALS AND METHODS

### Experimental Design

This study investigated intrahepatic determinants of clinical outcome in patients with BA through retrospective analysis and prospective sampling of BA and non-BA cholestatic control livers upon written informed consent of the patients’ parents. Human liver tissue and clinical data were obtained from patients undergoing KPE, diagnostic biopsy or LTX at Charité Universitätsmedizin Berlin, Hannover Medical School and University Children’s Hospital Tübingen. The study was approved by the Ethics Committee of Charité – Universitätsmedizin Berlin (EA2/148/22) and conducted in accordance with the Declaration of Helsinki.(*44*)

After collection of liver tissue during LTX or KPE, samples were immediately snap frozen, stored in basal medium or in 10% Formalin at 4°C, respectively. Liver samples were further processed for respective experiments immediately or within a maximum of 24 hours after biosampling. Biospecimen collection and processing were reported in accordance with the applicable items of the Biospecimen Reporting for Improved Study Quality (BRISQ) guidelines.(*45*)

### Single-cell transcriptomics

For snRNA-seq, we used fresh frozen liver tissues from patients with BA and cholestatic controls (control n = 3, mild n = 4, severe n = 4). Nuclei were isolated from tissue pieces with less than 0.3 cm diameter and prepared for snRNA-seq capture as previously described.(*46*) In brief, tissues were homogenized in citric acid buffer (250 mM sucrose, 25 mM citric acid, 1 μg/ml Hoechst 33342) with glass tissue douncers. The homogenate was filtered through 35 μm cell strainers to remove debris, then centrifuged at 500g for 5 min at 4°C. Nuclei were washed in citric acid buffer and spun down again for a final wash before being resuspended in cold resuspension buffer (25 mM KCl, 3 mM MgCl_2_, 50 mM Tris-buffer, 0.4 U/μl RNaseIn (2313A, Takara), 1 mM dithiothreitol, 0.4 U/μl SuperaseIn (AM2694, Thermo Fisher Scientific) and 1 μg/ml Hoechst 33342). After removing the supernatant, nuclei were counted and immediately loaded into the 10x Chromium Controller using the 10x Genomics Single Cell 3′ Library Kit v3.1 (PN 1000223, PN 1000157, PN 1000213 and PN 1000122). Libraries were pooled and sequenced on the NovaSeq 6000 system using paired-end mode (Illumina).

Raw sequencing data were processed using Cell Ranger v7.2.0 (10x Genomics). Transcripts were aligned to the reference human genome GRCh38.p13. Ambient RNA removal was performed with cellbender v0.3.0 on all samples.(*47*) Pre-processing to remove low-quality cells was performed with Seurat v5.1.0 based on the following criteria: (i) >200 and <7500-9000 genes, (ii) <10% mitochondrial reads. After log-normalization and variable feature selection, samples were integrated with Harmony using the IntegrateLayers() function in Seurat.(*48*) Using the integrated layer, neighbors and clusters were calculated, PCA was run, and finally UMAP calculation. Doublet detection was performed for each sample using DoubletFinder 2.0.3(*49*) and removed from the dataset. Cell type assignment was performed by using known marker genes.(*50–53*) Pair-wise compositional changes in cellular neighborhood densities were calculated with MILO v1.10.0.(*54*)

To perform unsupervised trajectory analysis, scVelo v0.3.2 was used as it utilizes velocity-based models to infer the dynamics of gene expression based on RNA splicing ratios. Loom files were generated with velocyto v0.17.17.(*55*) Potential cell-cell interactions between the different cell types were identified using CellChat v2.1.2.(*56*) Differential interactions between patient groups were calculated using mergeCellChat() and compareInteractions() functions. “Interaction strength” refers to the communication probability of each interaction in the computed network, while “information flow” denotes the overall communication probability of a given pathway, calculated as the sum of probabilities among all interacting cell groups. GO term enrichment was performed with clusterProfiler v4.14.6.(*57*) Differentially expressed genes used as input had adjusted p-values < 0.05. Gene set scores were calculated using the function AddModuleScore() from Seurat. The interferon response score gene set is composed of the union of genes belonging to the GO Biological Process terms GOBP RESPONSE TO TYPE I INTERFERON (GO:0034340) and GOBP RESPONSE TO TYPE II INTERFERON (GO:0034341) but filtered to genes detected in the datasets.

### Generation of FLOs and conditioned media induced injury-modeling

FLOs were generated from induced pluripotent stem cells (iPSCs) as previously described.(*21*) On day 19 of differentiation, FLOs were treated with 40 µg/mL polyI:C (Miltenyi) or vehicle control (NaCl) for 22 hours. On day 20, medium was replaced with 600 µL of basal FLO medium, followed by addition of 600 µL of liver CM of BA or non-BA explants. FLOs were cultured under the respective medium condition for 5 or 7 days, with the medium replaced every two days, before fixation (30 min in 10% formalin at RT) and processing for mpIF.

### Isolation and culture of liver lymphatic endothelial cells

Liver tissue with or without capsule was digested into single cells. Cells were plated at a density of 1 × 10⁶ cells/cm² on collagen-coated plates (5 µg/cm²; rat tail collagen, Corning) and cultured in EGM-2 medium (Lonza) supplemented with Y-27632 (10µM) and 25 ng/mL VEGF-C. The following day, plates were washed three times with DPBS containing 1% P/S, and fresh medium was added. Cells were cultured until approximately 80% confluency was reached, typically after 7–10 days, before MACS sorting was performed. PDPN-positive cells were isolated by indirect magnetic cell sorting using mouse IgG anti-human PDPN antibody at a dilution of 1:400 for 30 min, followed by anti-mouse IgG magnetic beads according to the manufacturer’s instructions.

### Statistics

Differential gene expression testing was calculated with the Seurat FindMarkers() function using the Wilcoxon Rank Sum test, with significance set at p-value <.05 and log-fold change of 0.25. For multiple testing, p-values were adjusted using the Benjamini-Hochberg method and considered significant at adjusted p-value <.05. CellChat identified enriched pathways to specific conditions by performing a paired Wilcoxon test within the rankNet() function. ClusterProfiler applies a hypergeometric test for enrichment analysis, with significance thresholds set at Benjamini-Hochberg adjusted p-value <.05. Gene set score comparisons were calculated with a Wilcoxon Rank Sum test and adjusted with Benjamini-Hochberg method, significance set at adjusted p-value <.05. Significance was only reported if the calculated Cohen’s effect size *r* was classified as small, moderate, or large. For all non-sequencing assays one-way ANOVA or unpaired t-test were performed and statistical results were reported when considered significant at adjusted p-value <.05. Data for mpIF, clinical characteristics and qPCR are presented as individual data points, with the mean±SD indicated, unless otherwise stated.

## List of Supplementary Material

Materials and Methods

Fig S1 to S9

Data files S1 to S2 (Supplementary Material Antibodies; Supplementary Material Primers)

Movies S1 to S3 (3D whole-mount IF imaging)

References (*58-73*)

## Acknowledgment

MR, LS, RLC are grateful for support by Eva Luise und Horst Köhler Stiftung (Alliance4Rare Program). MR also acknowledges support by the Emmy Noether Programme of the German Research Foundation (DFG). Graphical abstracts and schemes were created with BioRender (https://BioRender.com). ChatGPT and Claude were used only for language improvement, not authorative data generation or interpretation.

## Funding

Alliance4Rare / Eva Luise und Horst Köhler Stiftung (LS, RLC, MR)

German Research Foundation (DFG) Collaborative Research Center/Transregio 412/1 grant 535081457 (MR)

German Research Foundation (DFG) Emmy Noether Programme grant RE 3749/2-1 (MR)

Berlin Institute of Health at Charité Medical Scientist Program (RLC)

German Research Foundation Project-ID 556479455 and SFB1382; Project-ID 403224013 (AG)

Federal Ministry of Education and Research (BMBF), partner of the ERDERA, European Union’s Horizon Europe research and innovation program grant N°101156595 (AG)

Lymphatic Malformation Institute and European Union ERC, PREVENT, 101078827; ERC, 3D-SpatiOmics, 101189497 (RH)

## Author contributions

Conceptualization: MR, PB, CC, LS, RLC

Methodology: MR, LS, RLC, MM, SQ, JW

Investigation: LS, RLC, MZK, MM, SQ, JW, DG, RD, RB, MK

Formal analysis: LS, RLC, MZK, MZ, AS, JL, AG

Visualization: LS, RLC

Resources: MU, JK, CS, OMS, BG, RÖ, JCK, SH

Project administration: LS, RD

Supervision: SE, RH, RE, PB, CC, MR

Writing – original draft: LS, RLC, MR, PB

Writing – review & editing: All authors.

All authors interpreted the data and approved the final version of the manuscript.

## Data and materials availability

The snRNA-seq and spatial transcriptomics sequencing data generated in this study have been deposited in the European Genome–Phenome Archive under the accession number EGAXXXXX. The data are available under controlled access due to the sensitive nature of sequencing data, and access can be obtained by contacting the data access committee of the study. The fully annotated single-cell and spatial transcriptomics objects are available at figshare (XXXXX) and can be easily explored through our interactive data explorer (XXXXX).

The mass spectrometry proteomics data have been deposited to the ProteomeXchange Consortium via the PRIDE partner repository with the dataset identifier PXD085983. Reviewers can log in to the PRIDE website using the following details: Project accession: PXD085983, Token: 6iQrdQsZSNjq.

