## Supplementary Figures, Material and Methods for "Lymphatic Dysplasia Shapes Liver Epithelial Outcomes in Biliary Atresia"

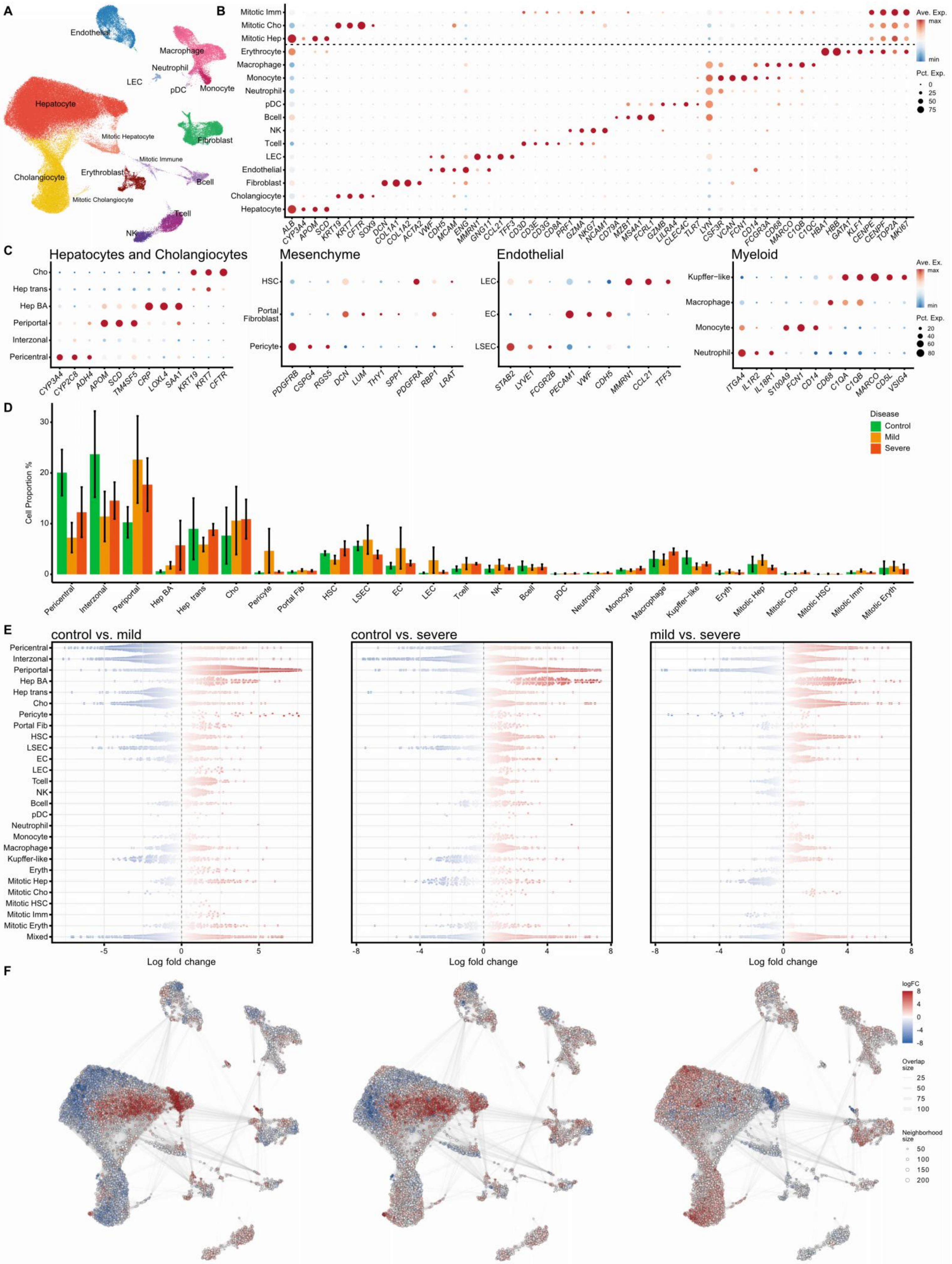


**Fig.S1 Single-nucleus RNA sequencing detects frequent and infrequent cell types with nuanced**

**abundance across patient groups. A+B:** General annotation and markers used to identify broad cell types. **C:** Cell specific markers differentiating cell type identities. **D:** Proportion of different cell clusters across disease severities, mean ± SEM. **E:** Compositional log fold changes of cellular neighborhoods between conditions. **F:** UMAP of log fold changes of each cellular neighborhood per comparison. Dot size depicts number of cells per neighborhood.

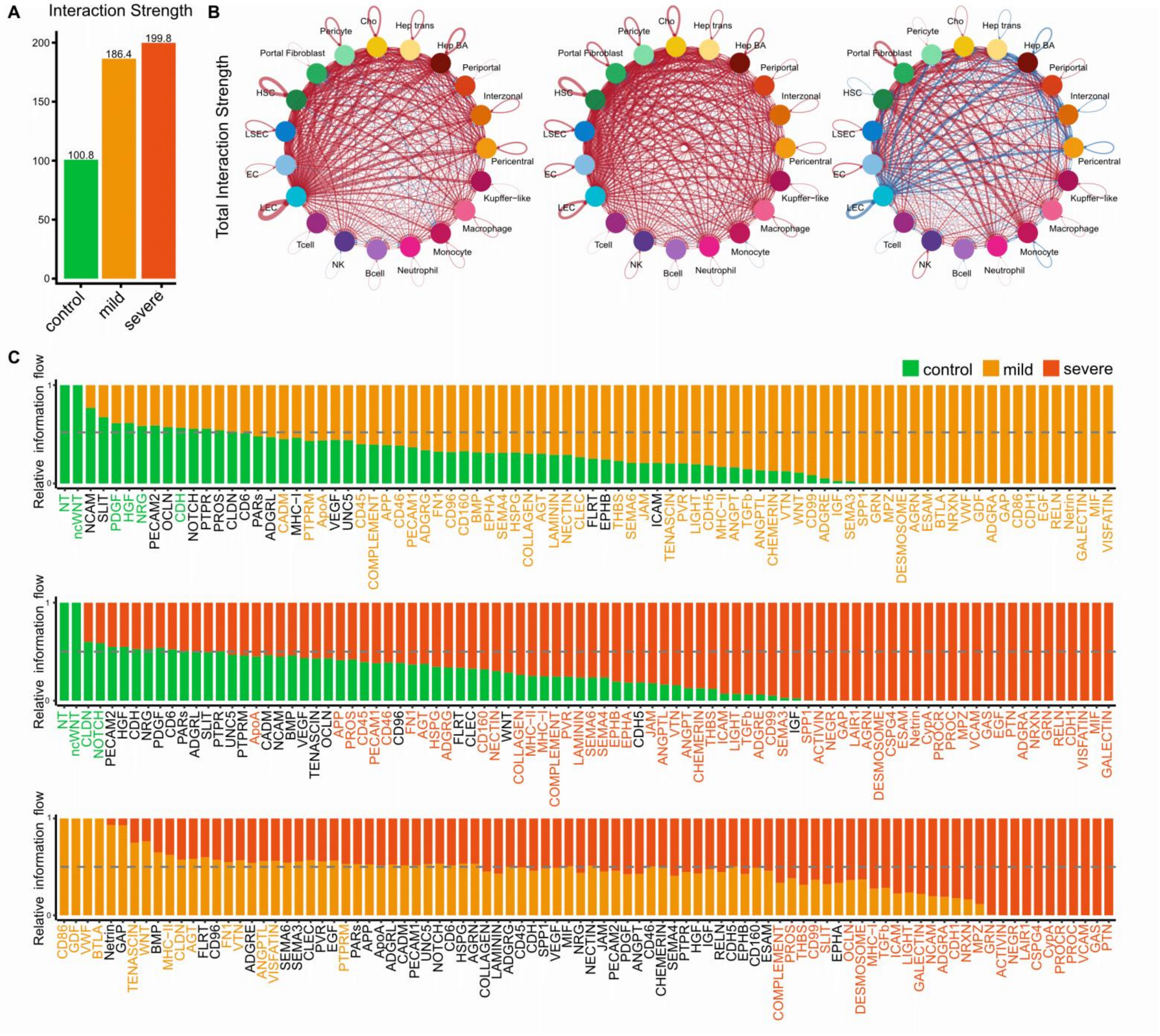

**Fig.S2 Distinct inferred interactions in mild versus severe BA. A:** Cell-cell interaction inference analysis for interaction strength in cholestatic control, mild and severe BA patients. **B:** Chord diagrams show inferred interaction count and strength between cholestatic control, mild BA and severe BA patients. **C:** Ranked interaction pathway enrichments between cholestatic controls, mild BA and severe BA patients.

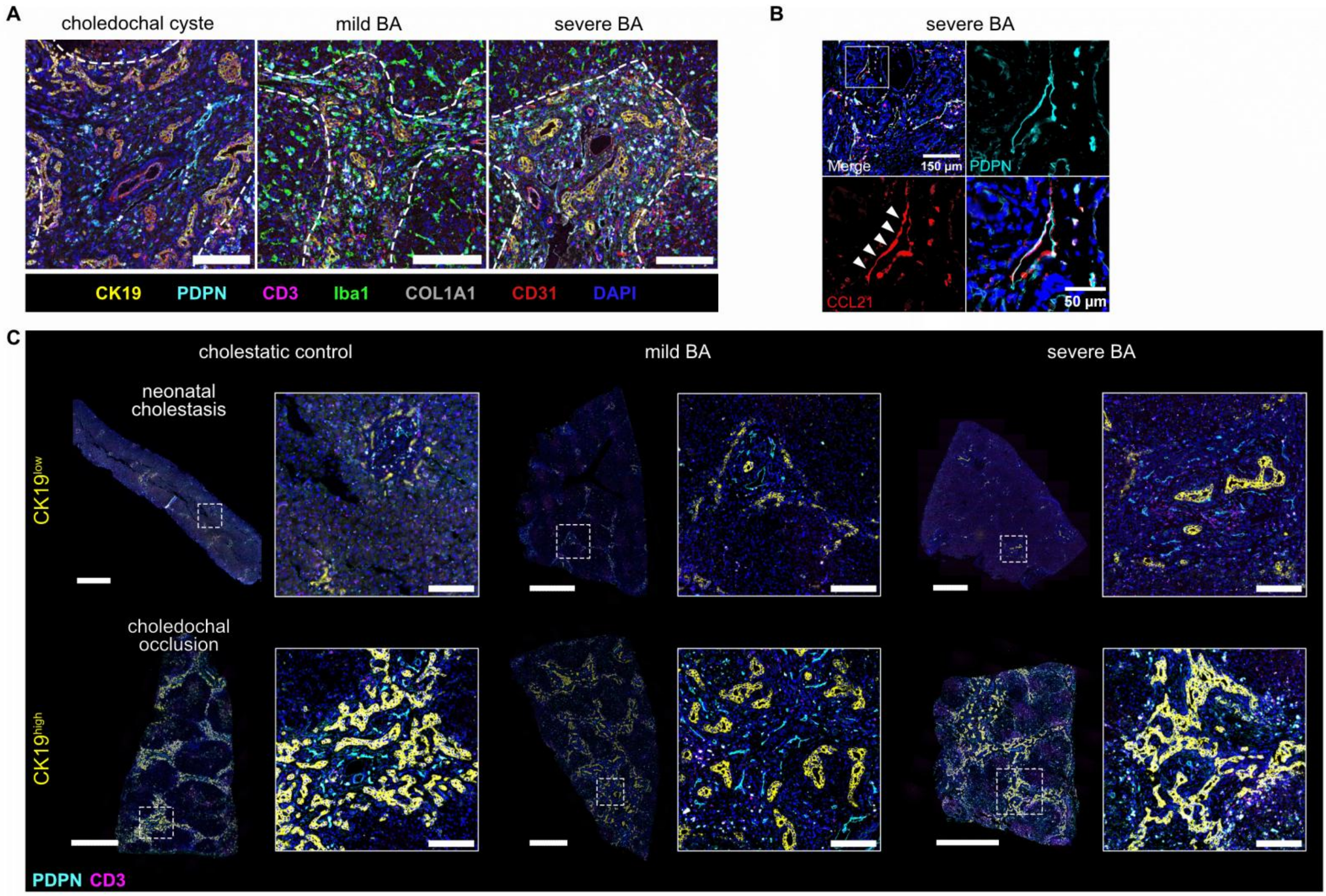


| | cholestatic control | mild BA | severe BA |
|---|---|---|---|
| CK19low | 6 (66.7%) | 9 (90%) | 2 (22.2%) |
| CK19high | 3 (33.3%) | 1 (10%) | 7 (77.8%) |

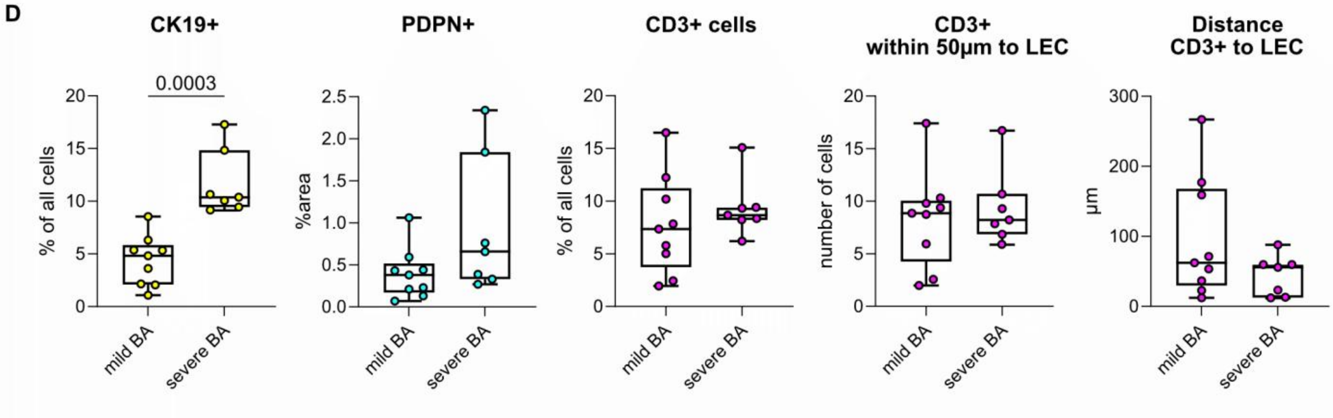


**Fig.S3: CCL21$^{high}$ and CK19$^{high}$ tissue signature marks severe BA. A:** Representative mpIF images of a cholestatic control, mild BA and severe BA patient show portal area with ductular cells (CK19, yellow), LECs (PDPN, cyan blue), T cells (CD3, magenta), macrophages (Iba1, green), fibrotic tissue (COL1A1, grey), vascular endothelium (CD31, red) and nuclei (DAPI, blue). Scale bars: 150µm. **B:** Representative mpIF image of severe BA shows LECs (PDPN, cyan blue), CCL21 (CCL21, red) and nuclei (DAPI, blue) in portal areas.

**C:** Representative mpIF images of complete tissue sections and zoom in of CK19$^{low}$ and CK19$^{high}$ liver samples show ductular cells (CK19, yellow), LECs (PDPN, cyan blue) and T cells (CD3, magenta). Scale bars in full tissue images: 1000 µm. Scale bars in zoom in: 150 µm. Table shows number of patients per group with CK19$^{low}$ or CK19$^{high}$ tissue signature. **D:** Boxplots show minimum to maximum and interquartile range of %CK19+ cells, %PDPN+ area, %CD3+ cells, number of CD3+ cells within 50µm around LECs and mean distance of CD3+ cells to LECs in µm for CK19$^{low}$ mild versus CK19$^{high}$ severe BA subgroup analysis. Unpaired t-test results are only indicated if $p<0.05$.

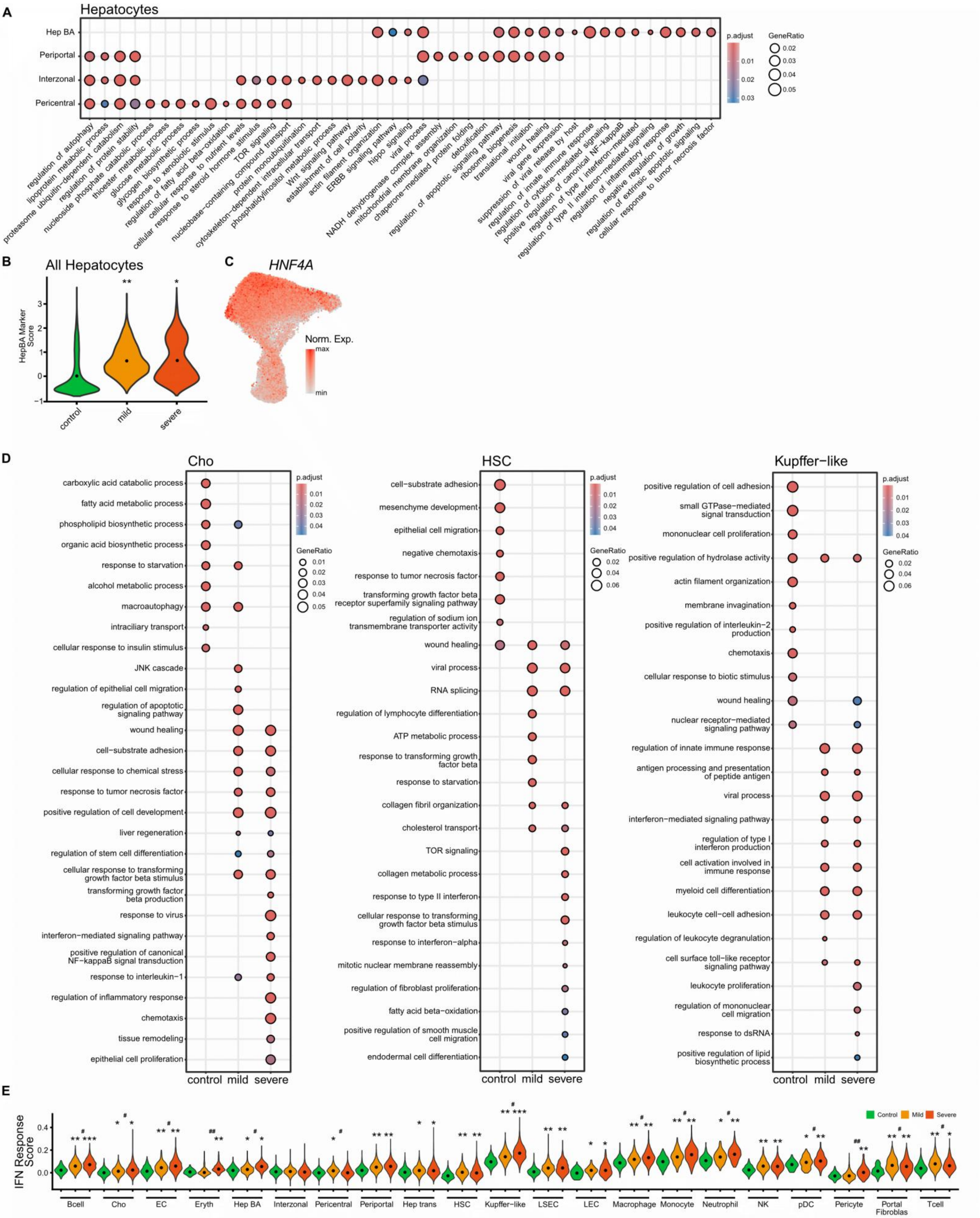


**Fig.S4 Functional characterization of major cell types in BA and cholestatic controls reveals distinct programs in severe BA. A:** Top GO terms across hepatocytes. **B:** Hep BA gene signature score measured for all hepatocytes. Dot indicates median scores per condition. Significance is indicated with a * (control vs. mild/severe) or # (mild vs. severe) for comparisons that reached significance and notable effect sizes: */# - small, **/## - medium, ***/### - large. **C:** HNF4A expression in hepatocytes and cholangiocytes. **D:** Top GO terms across disease conditions for cholangiocytes, hepatic stellate cells (HSC) and Kupffer-like cells. **E:** Interferon response score across all cell types and disease conditions. Dot indicates median scores per condition. Significance is indicated with a * (control vs. mild/severe) or # (mild vs. severe) for comparisons that reached significance and notable effect sizes: */# - small, **/## - medium, ***/### - large.

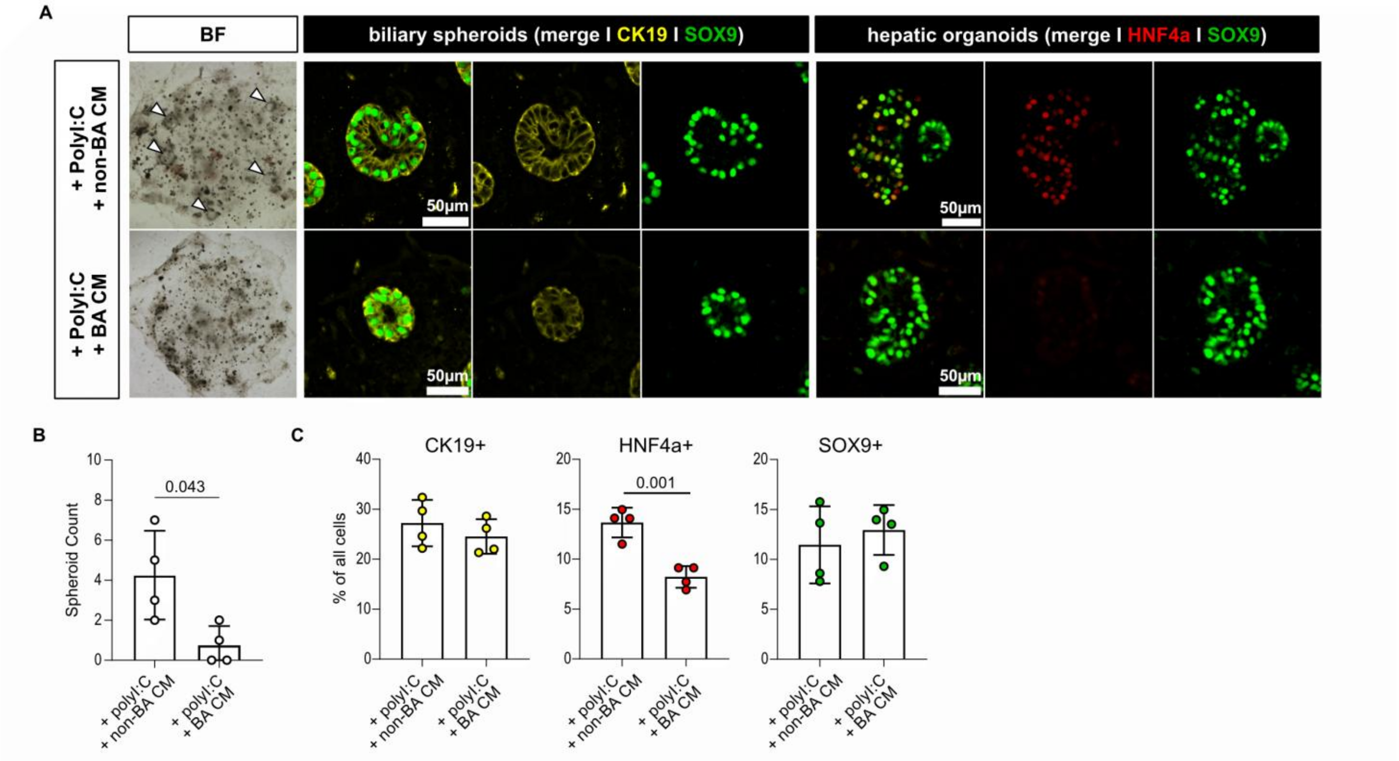


**Fig.S5 BA-specific HNF4A loss in a developmental liver-like organoid model. A:** Bright Field (BF) and immunofluorescence (IF) images of FLOs after polyI:C + non-BA CM and polyI:C + BA CM treatment. **B:** Bar graphs show mean±SD for number of spheroids per dome. **C:** Bar graphs show mean±SD for % of CK19+, HNF4a+ and SOX9+ cells respectively. Unpaired t-test results are only indicated if p<0.05.

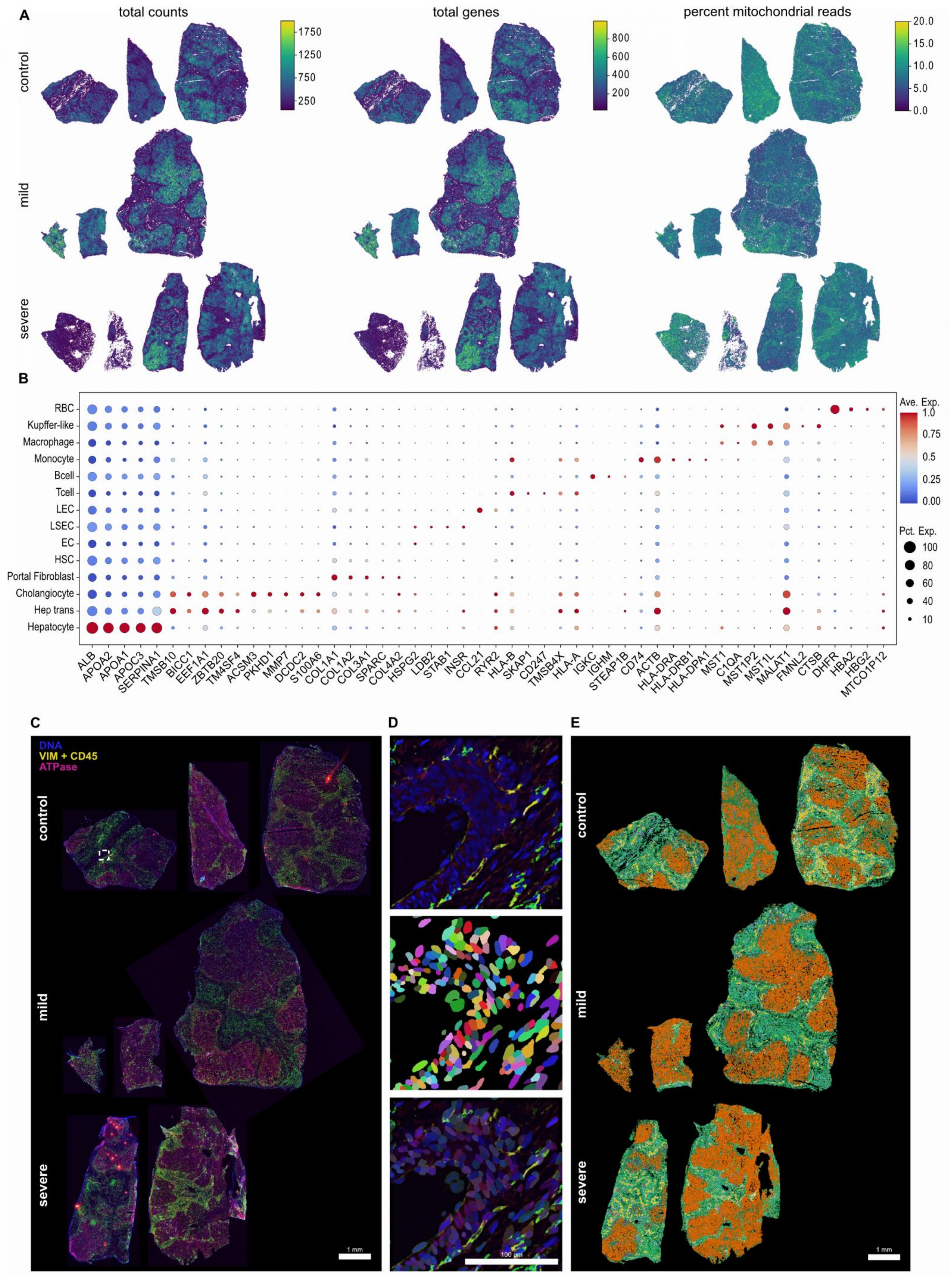


**Fig.S6 Stereo-seq spatially resolves the cell types identified by snRNA-seq.**

**A:** Spatial distribution of total UMI counts, total genes and percent mitochondrial reads in all samples. **B:** Top 5 differentially expressed genes in the identified cell types. **C:** Multiplex immunofluorescent (mIF) staining for VIM+CD45 and ATPase in all tissues that was used for cell segmentation. Scale bar = 150 µm. **D:** Preview of the cell segmentation output of the retrained µSAM model. Bounded box zoom in from Fig.S6C. Top: mIF preview, Middle: µSAM cell segmentation, Bottom: overlay. Scale bar = 100µm. **E:** Spatial distribution of identified cell types in liver tissues from all samples.

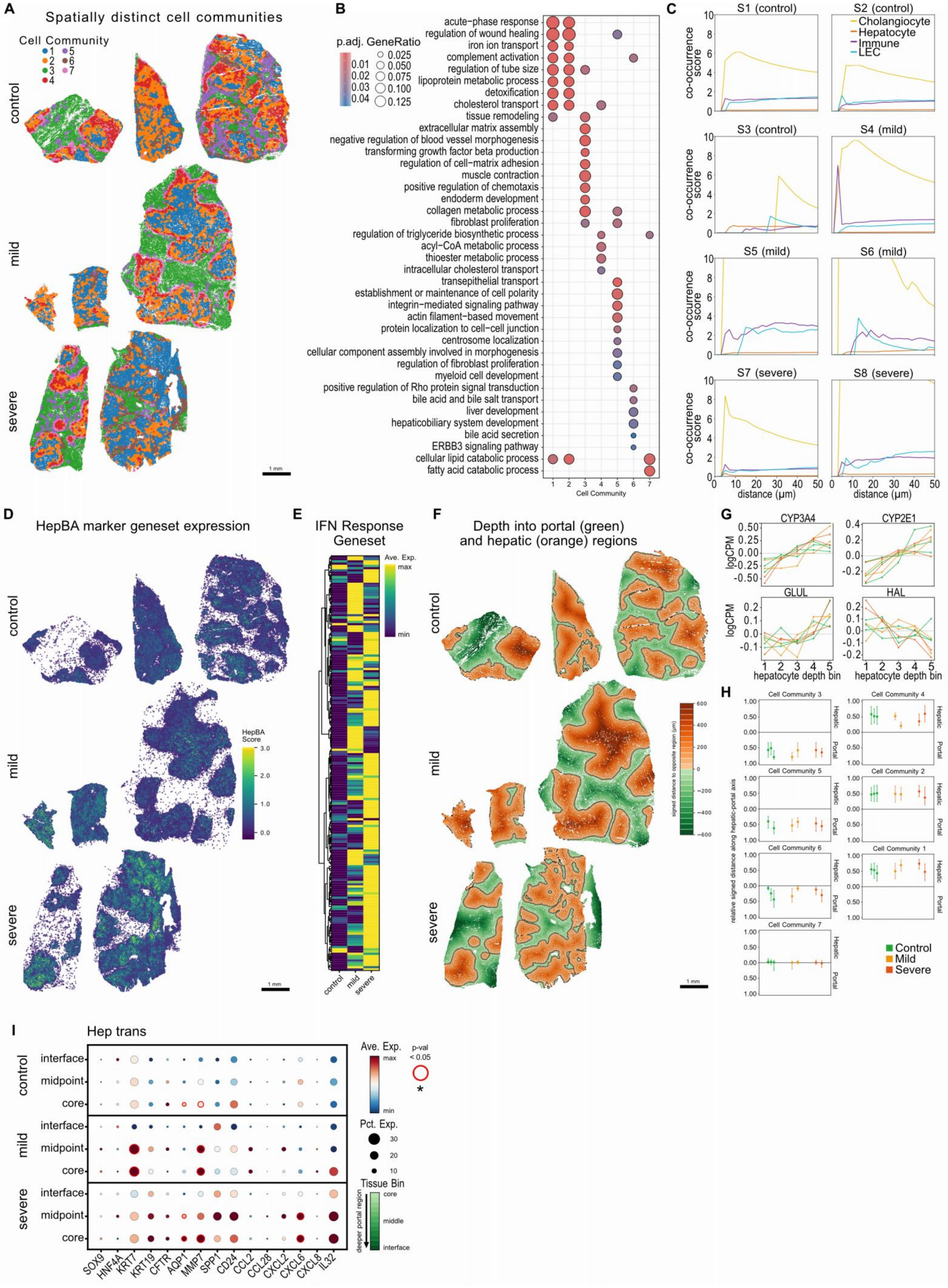

**Fig.S7 Spatial characterization of cellular communities and phenotypes.**

**A:** Spatial distribution of identified cellular communities in all samples. Scale bar = 1mm. **B:** Unique GO terms enriched in the cellular communities. **C:** Cellular co-occurrence analysis of hepatocytes, LECs, and immune cells relative to cholangiocytes in all samples. **D:** Average gene expression of interferon response genes between conditions. **E:** Spatial distribution of HepBA geneset score expression in all samples. **F:** Signed distance measurement of depth relative to hepatic and portal regions. Thick border represents the interface between the two regions. Concentric rings represent 100µm bins within region. Maximum depth is 600µm from the interface. **G:** Expression of pericentral and periportal zonation markers along the hepatocyte region bins in logCPM for each sample. **H:** Distribution of identified cellular communities across hepatic or portal regions. Dots represent the mean and the whiskers represent the IQR. **I:** Expression of ductular reaction and inflammatory markers in cholangiocytes when binning the regional depth into 3: interface, midpoint and core. Significant differences are depicted by red circle outlines if a midpoint or core regions are significantly regulated compared with interface and an asterisk if a significant difference is noted between midpoint and core regions.

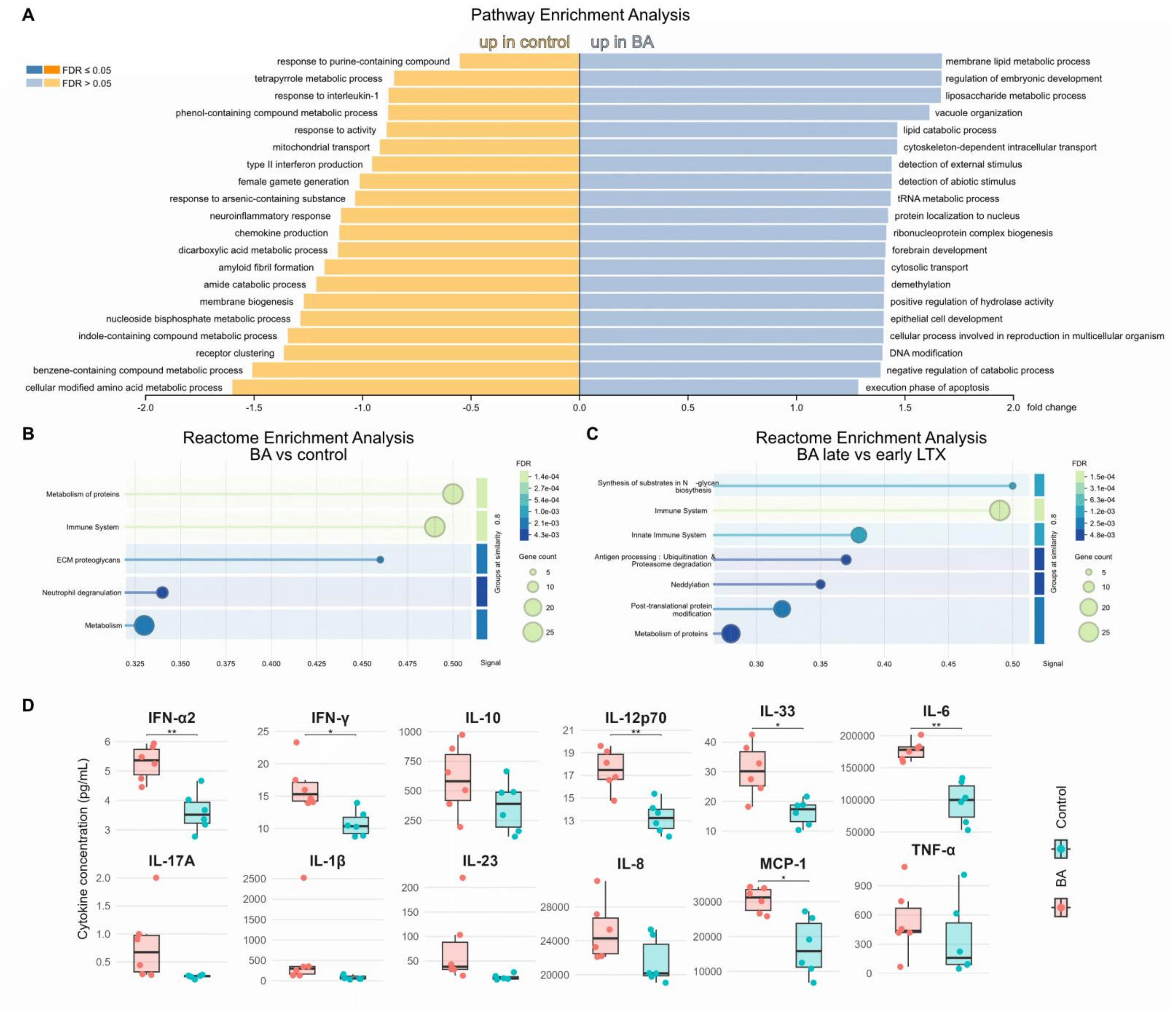


**Fig.S8 In-depth profiling of liver conditioned media reveals unique soluble mediators in severe BA. A:** Pathway Enrichment Analysis for BA versus non-BA CM from proteomic findings. **B:** Reactome Enrichment Analysis for BA versus non-BA CM from mass spectrometry findings. **C:** Reactome Enrichment Analysis for BA CM from 'severe' (early LTX) versus 'mild' (late LTX) BA patients. **D:** Boxplots show median±IQR for cytokines profiled from BA and non-BA CM by LegendPlex Assay. *<.05; **<.01.

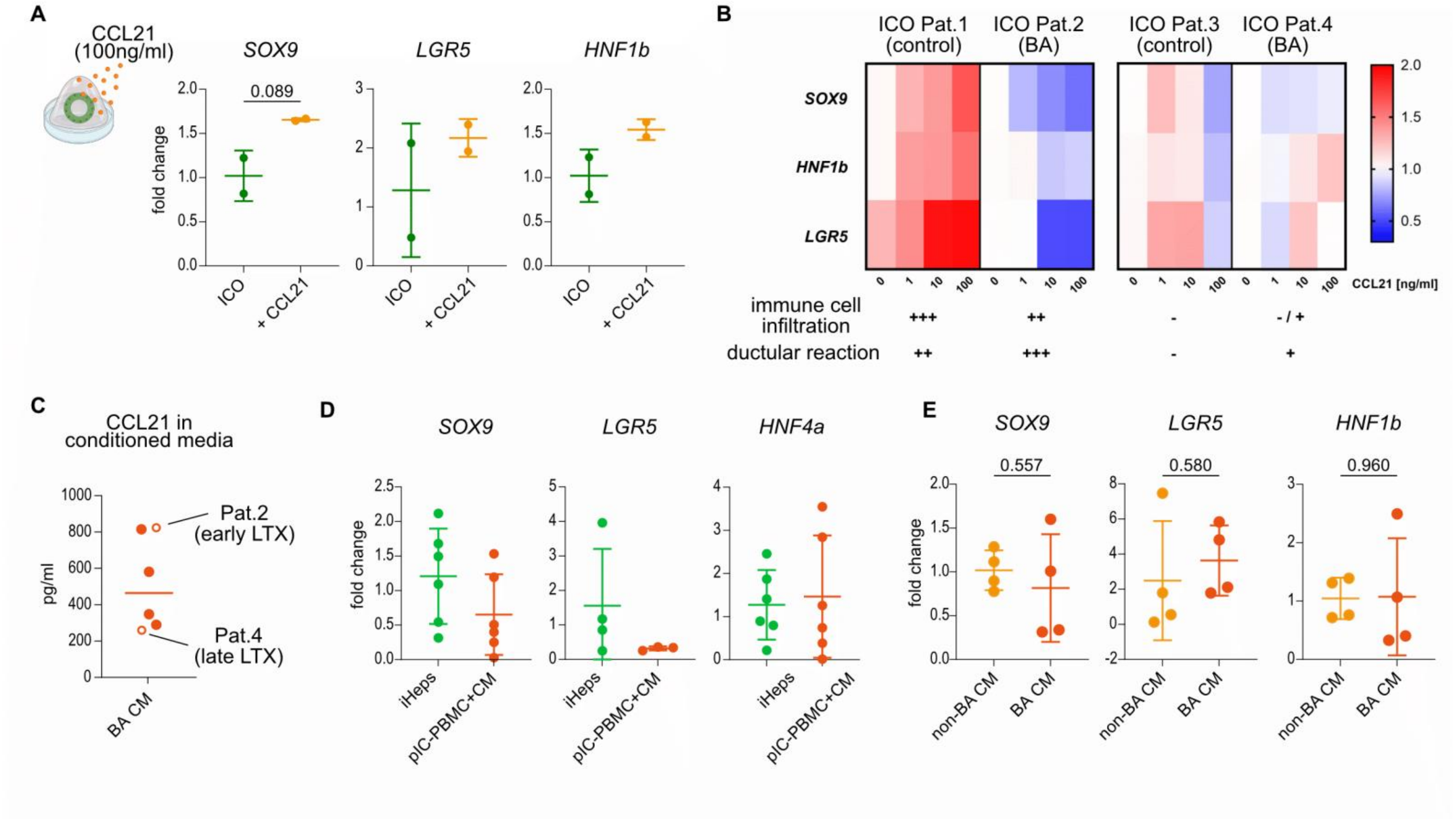


**Fig.S9 CCL21-sensing is dependent on inflammatory priming in an immune cell rich environment. A:** CCL21 treatment (100ng/ml) of ICOs. Graphs show mean±SD of fold change up-/downregulation of respective genes. **B:** Heatmap of fold change values of SOX9, HNF1b and LGR5 acquired by rt-qPCR of four different ICO lines treated with CCL21 (1, 10 and 100ng/ml). Grading (- / + / ++ / +++) for level of immune cell infiltration and ductular reaction was done based on histological findings in IHC or IF liver stainings of the resepective ICO-donor. **C:** Levels of CCL21 in liver CM shown for six individual BA patients and acquired by ELISA. **D:** Treatment of iPSC-derived hepatocytes with BA CM and pI:C-primed PBMCs. Graphs show mean±SD of fold changes of respective genes. **E:** Treatment of ICOs with BA and non-BA CM. Graphs show mean±SD of fold change up-/downregulation of respective genes. Unpaired t-test results are only indicated if $p<0.05$.

### Supplementary materials and methods

#### Stereo-mpIF-seq spatial transcriptomics

For spatial transcriptomics, we used matched fresh frozen tissues that were also used for snRNA-seq. Multiple tissue pieces were embedded in Tissue-Tek OCT Compound (4583, Sakura) inside a 1cm2 cryomold (4565, Sakura) and allowed to solidify on dry-ice. Using a cryotome (CM1950, Leica), 10 µm-thick tissue sections were made and loaded onto Stereo-seq T-chips v1.2 (211ST114, STOmics). The succeeding steps of tissue fixation, antibody staining, fluorescence microscopy, tissue permeabilization, reverse transcription, tissue removal, cDNA release and library generation were all performed according to the manufacturer's instructions unless otherwise specified below (Manual Version A, Mar 2023, STOmics). The antibody panel used were Anti-Vimentin (1:250, M7020, Dako), Anti-CD45-AF555 (1:250, 62267, Cell Signaling), Anti-Sodium Potassium ATPase (1:500, ab76020, Abcam), Goat anti-Mouse AF488 (1:1000, ThermoFisher, A32723) and Goat anti-Rabbit AF750 (1:1000, ThermoFisher, A21039). To prevent cross-reactivity, primary antibodies against Vimentin and Sodium Potassium ATPase were incubated for 25 minutes, followed by the secondary antibodies for 10 minutes, and finally the CD45-AF555 for another 25 minutes for a total staining time of 1 hour. Stained tissues were imaged with a slide scanning fluorescence microscope (Metafer, MetaSystems). Tissues were permeabilized for 10 minutes with 1X Permeabilization Reagent Solution. Libraries were sequenced on the DNBSEQ-T10 system using paired-end mode (MGI).

Raw sequencing data were processed using SAW v8.1 (STOmics). Transcripts were aligned to the reference human genome GRCh38.p13. To improve transcript assignment to cells, the vision foundation model for image segmentation, µSAM, was used to identify cellular boundaries of the tissues using the mpIF images of the Stereo-seq chips.*(58)* Due to the extreme autofluorescence of fibrotic tissues, the Vimentin channel was used perform autofluorescence removal on the CD45 channel using PCA background correction functions from scikit-image v0.25.2 and scikit-learn v1.7.0. A final RGB image of the DAPI, CD45+VIM (PCA corrected) and Sodium Potassium ATPase were generated for each Stereo-seq chip. To increase cell segmentation performance on our images, μSAM v1.6.2 "vit_b_lm" model was retrained on 25 manually annotated images across all tissue regions and samples (20 – training, 5 – validation) on an NVIDIA RTX 6000 Ada 48GB GPU. The fine-tuned model was then used for cell segmentation of the mIF images. The resulting cell masks were then assigned all the transcripts whose spatial coordinates fall within the instances. This was achieved by following the "stain segmentation" methodology from Spateo v1.1.1, which outputs h5ad and GEM files for downstream use.*(59)*

The datasets were analyzed through scanpy v1.9.6, squidpy v1.2.2, and stereopy v1.2.2 depending on the workflow needed.*(60-62)* The samples were filtered following these criteria: (i) <50 and >1000-2000 counts, (ii) <5 and >500-1000 genes, (iii) >20% mitochondrial reads. Due to these thresholds, we identified 2 samples to be of low quality and were thus subsequently dropped from all downstream analysis. The dataset was normalized through Pearson residuals *(63)* and integrated with Harmony.*(48)* Finally, neighbors, clusters, principal components, and a UMAP were calculated. To leverage the use of the matched annotated snRNA-seq dataset, we inferred cell type composition using RCTD using "doublet mode".*(64)* Cell type annotations were generalized for the inference (i.e. hepatocytes vs. periportal, interzonal and pericentral hepatocytes; fibroblasts only vs. portal fibroblasts and HSCs). Due to the compartmentalized distribution of some cell types, these generalized identities were split if cells localize within the hepatic or portal region of the tissue using a manual tissue region mask based on the mpIF stainings. Fibroblasts were categorized as portal fibroblasts if within the portal region and HSCs if within the hepatic region. Endothelial cells were categorized as endothelial in the portal region and as LSECs in the hepatic region. Finally, macrophages were categorized as macrophages in the portal region and as Kupffer cells in the other. Conserved cellular communities across all samples were calculated with a predetermined 73x73µm window (~30-50 cells) relative to the tissue sizes using Stereopy's community_detection() algorithm with agglomerative clustering. Cellular co-occurrence was calculated with squidpy's co_occurrence() function using a specific cell type as the point of reference. To measure immune infiltration at the portal area, all identified immune cells were grouped as "immune" for this analysis. Tissue region depth analysis was performed by using k-d tree nearest neighbor search implemented in scipy v1.10.1. Distances of cell were measured and signed according to the depth of the tissue with positive values for the hepatic region, negative for the portal region, while zero denotes the interface. The manual tissue region mask was used for this analysis. Cells were divided into five or three equally sized bins according to their distances to the opposite region. For pseudobulks, raw UMI counts were summed across cells within each sample, normalized to counts per million and log-transformed. Hep BA gene set scores were calculated using the function score_genes() from scanpy on the top 50 differentially expressed genes of Hep BA cells from the snRNA-seq dataset.

**Multiplex immunofluorescence imaging**

Sequential multiplex immunofluorescence was performed based on the protocols previously described with small adjustments were necessary.*(65,66)* Briefly, antigens were retrieved using a citrate buffer (pH 6.0) and nonspecific antibody binding was blocked with 2% normal donkey serum. At the end of each antibody staining and imaging cycle, antibodies were eluted at 56°C for 1 hour in the antibody stripping buffer containing 62.5 mM Tris-HCl (pH 6.8), 2% (w/v) SDS and 114.4 mM β-mercaptoethanol prepared in deionized water. Images were registered and prepared by using the CytoPrixm software as described previously.*(65)* CytoPrixm can be freely downloaded at https://zenodo.org/records/19059527.

**Whole mount immunofluorescence and light sheet microscopy**

Whole mount IF was performed as previously reported.*(67)* Briefly, snap-frozen liver pieces of a maximum of 2 x 2 x 2 mm were fixed, permeabilized and blocked prior to IF staining for PDPN (Santa Cruz, 376695), CK19 (R&D, AF3506) and CD45 (rabbit, 19581). After stepwise dehydration, samples were optically cleared in a benzyl alcohol:benzyl benzoate solution. Liver samples were then imaged on a LaVision Ultramicroscope II (LaVision BioTec). 3D reconstruction was performed using Imaris (version 9.8, Oxford Instruments Abingdon, UK).

**Generation and culture of human intrahepatic cholangiocyte organoids**

Human ICOs were generated following an adapted protocol from the adult human liver organoid culture method described by Huch et al. and Broutier et al..*(68,69)* Briefly, human liver tissue samples obtained from explanted livers or from KPE specimens were mechanically minced and enzymatically digested for up to 90 minutes at 37°C in a digestion buffer containing collagenase type IV (2.5 mg/ml, Worthington Biochemical Corporation, distributed by CellSystems GmbH; #LS004188) and DNase I (0.1 mg/mL; Roche; #10104159001). Digestion was then terminated by the addition of Wash Medium (WM), and the cell suspension was washed several times with WM and ice-cold Dulbecco's phosphate-buffered saline (DPBS; Gibco; #14190136) before being passed through a 70-µm nylon cell strainer and seeded at a density of approximately 0.5–1 × $10^6$ cells per 50-µL Matrigel (Corning, #CLS356231) droplet.

Following polymerization of the Matrigel, newly established cultures were maintained for 3 days in liver initiation medium (LIM) under standard conditions (37°C, 5% $CO_2$; concentrations and suppliers in Supplementary Table MK1). Cultures were subsequently maintained in liver expansion medium (LEM), with medium being replaced every 2 days, and organoids were mechanically passaged approximately every 2 weeks at a split ratio of 1:3 or 1:4. For subsequent experiments, cultures between passages 4 and 7 were employed.

**Co-culture of LECs and ICOs**

For the co-culture with ICOs, two wells of ICOs (80% confluency) were pooled into one well of an AggreWell400 plate (STEMCELL Technologies), either with or without LECs. LECs were added at 8,750 or 45,000 cells per well according to cell availability using co-culture medium supplemented with Y-27632. The following day, the contents of each AggreWell were transferred 1:2 or 1:3 to a 24-well plate containing 50 µL of Matrigel per well. Co-culture medium consisted of LEM basal medium supplemented with the full concentrations of growth factors from both EGM-2 and LEM media, together with 25 ng/mL VEGF-C; or EGM-2 medium and complete LEM medium mixed at a 1:1 ratio, with VEGF-C added to a final concentration of 25 ng/mL after optimization. The medium was replaced every other day. On day 9, EpCAM-positive cells were selected using CD326 MicroBeads (Miltenyi Biotec).

**CCL21 treatment and bright-field imaging**

ICOs between passages 4 and 7 were mechanically passaged and reseeded in Matrigel as described above. Immediately following passaging, cultures were treated with recombinant human CCL21 (PeproTech; #300-35A-20) at final concentrations of 0.1, 1, 10, or 100 ng/mL in LEM, alongside a matched untreated control, and cultures were maintained for 7 days under standard conditions. Medium was replaced on days 2 and 4 with freshly prepared LEM containing the corresponding concentration of CCL21. Bright-field images were acquired using a Keyence BZ-X800 microscope. Whole-dome images were acquired using a 4× objective on days 1, 3, and 7 of culture, with day 1 serving as the baseline measurement. On day 7, the culture medium was aspirated, and the organoids were harvested by resuspending in TRIzol reagent (Invitrogen/ThermoFisher Scientific, # 12044977) and stored at −20°C until further analysis.

**Human iPSC-derived hepatocytes**

For differentiation of hepatocyte-like cells we utilized the human iPSC lines BIHi001-B, BIHi005-A and BIHi001-A provided by the BIH Core Unit Pluripotent Stem Cells and

Organoids. For maintenance, we kept all hiPSCs in mTeSR medium (Stem Cell Tech), cultured on vitronectin coated plates.

We adapted the 28-day protocol described in Weihs et al.,*(20)* adding EMT inhibitors described in Li et al from day 13 on (HBM Stage) until the end of the differentiation.*(70)* In Brief, we first adapted hiPSCs in mTeSR medium to 10 µM ROCK-inhibitor for at least 5 days. We then initiated the hepatic differentiation by single-cell splitting on Geltrex-coated plates with the following strategy for daily medium changes:

Days 0–5: DE-induction medium (RPMI1640, 2% B27, 1% PenStrep, 100 ng/ml Activin A, 50 ng/ml Wnt3, 0.5 mM sodium butyrate), followed by one day without sodium butyrate.

Days 6–12: KO-medium (KO-DMEM, 20% KO Serum, 1% PenStrep, 1% Glutamax, 1% NEAA, 1% DMSO, 0.1 mM β-mercaptoethanol) for hepatic commitment.

Days 13–19: HBM medium (Lonza) supplemented with 20 ng/ml HGF, 20 ng/ml OSM, and 1% PenStrep including 10 µM SB431542, 20 µM Forskolin, 0.5 µM IWP2, 5 µM DAPT and 0.1 µM LDN193189.

Days 20–28: Maturation in HCM Bulletkit medium (Lonza, excluding EGF), with supplementation of 5 nM T3 (Tocris), 100 nM sodium selenite (Sigma), 100 µM CDCA (Sigma), 10 µM SB431542, 20 µM Forskolin, 0.5 µM IWP2, 5 µM DAPT and 0.1 µM LDN193189.

**Isolation of human peripheral blood mononuclear cells (PBMCs)**

Blood isolation was done in accordance with ethical standards of the Charité - Universitätsmedizin Berlin and the amended Declaration of Helsinki. We isolated PBMCs from a healthy individual who was assessed by a physician to ensure the absence of clinical illness or medication intake. Specifically, we mixed whole blood with PBS (w/o Ca2+ Mg2+) in a 1:1 ratio and overlaid this dilution on the Ficoll in a 3 to 2 ratio, respectively. We then centrifuged the mixture for 30 minutes at 300 g at room temperature, using the lowest levels of brake and acceleration. The leukocyte layer was transferred to a new tube and washed once with PBS. After lysing red blood cells by incubating the cells for 10 min in ACK lysis buffer, we washed twice, counted and froze cells down in a 10% DMSO solution in FCS using Mr. Frosty. The cells were stored in vapor phase liquid nitrogen below -135°C until further use.

**Modelling the BA environment on hepatocytes**

For direct CCL21 exposure, we added the recombinant protein or vehicle to the medium from day 24 on for 5 days, using the indicated concentrations with daily medium changes. 500 µM or 200 µM CDCA (Sigma) was added in the respective conditions.

For PBMC priming, we thawed the cells 18 h before co-culture and maintained them in RPMI medium containing 10% heat-inactivated fetal calf serum (hiFCS) and Pen-strep (~10^6 cells/ml). Within this 18 h pre-incubation, PBMCs incubated either with 5 µg/ml lipopolysaccharide (LPS, eBioscience or Stem Cell Technologies) or 40 µg/mL polyI:C. Before adding them to the inner compartments of 0.4 µm pore size millicell trans-well inserts (Millipore; PBMCs:Hepatocytes = 1:4) at day 27 of hepatocyte differentiation, we washed off the RPMI medium and re-suspended them either in pure hepatocyte medium, mixed 1:1 with BA CM, or pure hepatocyte medium supplemented with recombinant CCL21 for the respective hepatocyte treatment conditions. The treatment was continued for 48h during co-culture. All tested conditions had the same total volume of medium.

**RNA isolation, cDNA synthesis, and quantitative real-time PCR**

Total RNA was isolated from ICOs or hepatocytes using the Arcturus PicoPure RNA Isolation Kit (ThermoFisher Scientific; #12204-01) according to the 'turer's instructions. Briefly, 100 µL chloroform was added to each sample per 500 µL TRIzol reagent; samples were then mixed and centrifuged at 12,000 × g for 10 min at 4°C. Subsequently, the aqueous phase was transferred to a fresh tube and mixed at a 1:1 ratio with 70% ethanol before being loaded onto RNA purification columns. Subsequent washing and on-column DNase digestion steps were performed following the manufacturer's instructions before the RNA was eluted in 11 µL Elution Buffer. The total RNA concentration was measured using the NanoDrop One/OneC Microvolume UV-Vis Spectrophotometer (ThermoFisher Scientific) before the samples were stored at -80°C until further processing. RNA was reverse-transcribed into complementary DNA (cDNA) using the High-Capacity cDNA Reverse Transcription Kit (Thermo Fisher Scientific; #4368813), according to the manufacturer's instructions.

Quantitative real-time PCR was performed using PowerUp™ SYBR™ Green Master Mix (Applied Biosystems; #A25776) on a QuantStudio™ 3 Real-Time PCR System (Thermo Fisher Scientific), according to the manufacturer's instructions. Primer sequences are provided in Supplementary Table MK2. Relative gene expression was normalized to RPS18 expression and calculated using the $2^{-\Delta\Delta Ct}$ method, with the corresponding untreated control condition used as the calibrator.

**Generation of liver-conditioned medium**

Liver biopsies were freshly sampled and kept in DMEM/F12 supplemented with 10% FCS and 1% penicillin/streptomycin (P/S) for a maximum of 10 h until processing. Liver tissue was washed in PBS with P/S and minced thoroughly. 500 mg of minced tissue was transferred to 10 mL of basal medium (advanced DMEM/F12 with 1% HEPES, 1% GlutaMAX, 1% P/S, 1% N2, with or without 2% B27) and incubated for 22–32 h at 37 °C. For proteomic analyses, B27 supplementation was omitted. Conditioned medium was collected and centrifuged at 4,000 × g for 10 min to remove cellular debris, followed by sterile filtration through a 0.22-µm filter. The conditioned medium was aliquoted and stored at −80 °C until further use.

**Mass spectometry based bottom-up proteomics**

For proteomic analysis of liver-conditioned medium, 4.5 µl of B27-free liver-conditioned medium (n=7) was separated by one-dimensional SDS-polyacrylamide gel electrophoresis according to Beier et al..*(71)* Each sample lane was divided into eight fractions. In-gel digestion of proteins, peptide extraction, desalting, and sample preparation for Liquid Chromatography (LC)-Mass Spectometry (MS)/MS analysis were performed as described by Beier et al.*(71)*

LC-MS/MS analysis was performed using a nanoAQUITY UPLC system (Waters corporation, Milford, MA, USA) coupled to an LTQ Orbitrap Velos Pro mass spectrometer (Thermo Fisher Scientific Inc., Waltham, Massachusetts; USA). The LC gradient is described in Lerch et al..*(72)* Primary MS scans were acquired in the m/z range of 350-1750. MS parameters were taken from Beier et al.,*(71)* except for the exclusion time, which was set to 12 s.

MS/MS raw files were analyzed using MaxQuant (version 2.8.0.0, Max Planck Institute of Biochemistry, Martinsried, Germany, www.maxquant.org) according to the parameters described previously,*(71)* with N-terminal acetylation as an additional variable modification. Analysis was performed against protein FASTA databases of human proteins and isoforms (taxonomy id 3A9606_29, downloaded from UniProt on 27 June 2023), Bos taurus proteins (UniProt, 20 December 2023) and protein sequences of bovine serum albumin (BSA), bovine catalase, bovine peroxisomal catalase, and bovine superoxide dismutase.

Proteins were considered to be reliably identified if detected by at least two unique peptides, each with at least one MS/MS scan in two different samples or replicates, and if their total intensity divided by the number of unique peptides exceeded a mean of 100,000 across all replicates per sample. Protein quantification was performed using intensity-based absolute quantification (iBAQ) in MaxQuant.*(73)* Statistical significance was assessed using t-tests (p=0.01 and p=0.05) with Perseus 2.0.3.0 (Max Planck Institute of Biochemistry, Martinsried, Germany, www.maxquant.org). Proteins with more than 16-fold differences in iBAQ values were considered biologically different.

**LEGENDplex™ Assay**

Inflammatory cytokines and chemokines in explant liver CM from patients with BA and non-BA controls (n = 6) were quantified using the LEGENDplex™ Human Inflammation Panel 1 (BioLegend, #740809) according to the manufacturer's instructions. Undiluted samples were analyzed in duplicates using a 96-well filter-plate assay, alongside serially diluted standards and unconditioned medium controls. Acquisition was performed on a Cytek Aurora Full Spectrum Flow Cytometer (Cytek Biosciences, Fremont, CA, USA). Cytokine concentrations were calculated from analyte-specific standard curves using the LEGENDplex™ Data Analysis Software Suite (BioLegend, San Diego, CA, USA) and expressed in pg/mL following background correction against unconditioned medium controls.

### Supplementary Material: Antibodies

| Target | Host | Clonality | Conjugation | Application | Concentration | Manufacturer | Catalog number |
|---|---|---|---|---|---|---|---|
| CCL21 | goat | polyclonal | unconjugated | mpIF / neutralization | 1:20 / 0.2-1 µg/ml | R&D | #AF366 |
| CD3 | rabbit | monoclonal | unconjugated | mpIF | 1:1000 | abcam | ab16669 |
| CD31 | rabbit | polyclonal | unconjugated | mpIF | 1:200 | abcam | ab28364 |
| CD45 | mouse | monoclonal | unconjugated | mpIF | 1:400 | Agilent Dako | #M0701 |
| CD45 | rabbit | monoclonal | AF555 | whole-mount IF | 1:100 | Cell Signaling | #19581 |
| CK19 | sheep | polyclonal | unconjugated | mpIF / whole-mount IF | 1:400 / 1:100 | R&D | AF3506 |
| COL1A1 | rabbit | monoclonal | unconjugated | mpIF | 1:200 | Cell Signaling | 72026T |
| HNF4a | rabbit | monoclonal | unconjugated | mpIF | 1:100 | abcam | ab200142 |
| Iba1 | goat | polyclonal | unconjugated | mpIF | 1:200 | abcam | ab5076 |
| PDPN | mouse | monoclonal | unconjugated | mpIF / whole-mount IF | 1:400 / 1:100 | Santa Cruz | sc-376695 |
| SOX9 | rabbit | monoclonal | unconjugated | mpIF | 1:400 | abcam | ab185230 |

### Supplementary Material: Primers

| Gene | Forward primer (5'-3') | Reverse primer (5'-3') |
|---|---|---|
| CCR7 | CAACATCACCAGTAGCACCTGTG | TGCGGAACTTGACGCCGATGAA |
| HNF1B | TCACAGATACCAGCAGCATCAGT | GGGCATCACCAGGCTTGTA |
| HNF4A | ATCCAGGGAAGATCAAGCGGCTGC | CAGCATCTCCTGCAACAGGTTGTC |
| LGR5 | CCTGCTTGACTTTGAGGAAGACC | CCAGCCATCAAGCAGGTGTTCA |
| MMP7 | TCGGAGGAGATGCTCACTTCGA | GGATCAGAGGAATGTCCCATACC |
| SOX9 | TCCCCCAACGCCATCTTCAA | GGAAGTCGATAGGGGGCTGT |
